# Tick-Borne Pathogen Correlations with Primary Sclerosing Cholangitis: Ecological Evidence

**DOI:** 10.64898/2026.07.24.26358879

**Authors:** Kevin M. Johnson

## Abstract

**Background & Aims:** Primary sclerosing cholangitis (PSC) is a cholestatic liver disease of unknown etiology whose prevalence varies >30-fold worldwide, peaking in Northern Europe and the U.S. Upper Midwest. This geographic distribution is not fully explained by recognized risk factors. We examine its correlation with *Ixodes* tick exposure.

**Approach & Results:** PSC incidence across North America and Europe was compared with Lyme incidence, HLA-DRB1*03 frequency, latitude and other environmental factors. Autoimmune hepatitis (AIH) and primary biliary cholangitis (PBC) were included as controls. A U.S. analysis (MarketScan, 2018–2022; 110.7 million person-years) correlated age and sex-standardized rates against 24 exposures, including *Ixodes* density and tick-borne infections, using ancestry-adjusted partial correlations.

Cross-country PSC incidence tracked Lyme incidence (Spearman ρ = 0.71-0.87), a proxy for Ixodes exposure; HLA-DRB1*03, AIH, and PBC did not. Alaska Native and Greenlandic populations, high-latitude but without established human exposure to *Ixodes*-borne pathogens, report no PSC despite high autoimmune liver disease and inflammatory bowel disease. In the U.S., PSC was clustered and tracked *Ixodes*-borne pathogen incidence (ancestry-adjusted partial r, log scale: anaplasmosis +0.50, babesiosis +0.56, Powassan virus disease +0.52; in the Northeast–Midwest block, ancestry- and latitude-adjusted r = +0.72, +0.84, and +0.78, respectively). Non-*Ixodes* infections (*Ehrlichia chaffeensis* −0.26, spotted fever −0.40, tularemia −0.39), AIH, and PBC were null-to-negative; rural, agricultural, pollution, and healthcare-access also did not correlate.

**Conclusions:** These ecological data suggest the hypothesis that *Ixodes*-borne pathogen exposure is linked to PSC but cannot by themselves establish a causal role. Specifically, they raise the question whether such exposure is a trigger, occurring on a background of susceptibility. The findings are hypothesis-generating, yielding falsifiable predictions for case–control, serologic, and animal-model studies.

## Introduction

Primary sclerosing cholangitis (PSC) is a chronic, progressive cholestatic liver disease characterized by inflammatory and fibrotic destruction of the intrahepatic and extrahepatic bile ducts. It is strongly associated with inflammatory bowel disease (IBD), particularly ulcerative colitis, and frequently progresses to cirrhosis, cholangiocarcinoma, and liver transplantation.^1^

While the pathogenesis of PSC has been extensively studied, the fundamental question of what triggers PSC in the first place remains unanswered. Genetic studies have identified susceptibility loci, most prominently within the human leukocyte antigen (HLA) region, yet these explain only approximately 7–10% of disease liability.^2^ Published work often alludes to “environmental” factors, but little evidence for such factors has been found.^1,3^

An underexplored feature of PSC is its geographic distribution. Reported prevalence varies more than 30-fold worldwide, from among the highest levels in Northern Europe and the Upper Midwest United States to extremely low or zero prevalence in parts of Asia, the tropics, and the Arctic.^4–5^ Even within regions of similar ancestry and healthcare access, substantial differences exist: Finland has roughly twice the PSC incidence of some neighboring regions,^6^ and in the United States the Upper Midwest substantially exceeds Northern California.^7–8^ These gradients are difficult to reconcile with models based solely on genetics, diagnostic access, or generalized autoimmune susceptibility.

Populations in which PSC appears to be absent are also informative. Alaska Native populations have among the highest reported prevalences of autoimmune hepatitis (AIH) globally (42.9–58.6 per 100,000), yet population-based studies spanning decades have identified no cases of PSC.^9^ A survey from Greenland reports AIH and primary biliary cholangitis (PBC) but no PSC.^10^ These populations show that autoimmune susceptibility and northern latitude alone are insufficient to account for PSC; some additional element appears to be required.

Several epidemiologic features of PSC further distinguish it from most autoimmune diseases. PSC shows a consistent male predominance (approximately 60-65% of cases), whereas autoimmune diseases typically show female predominance.1 PSC has been reported to cluster in rural rather than urban environments,11 and case-control studies have reported increased exposure to farm animals.3 That framing requires qualification. The only case-control study of environmental exposures in autoimmune liver disease found place of residence (town versus countryside), housing type and well or spring water all null, with household pet contact the single positive association (odds ratio 2.6 to 3.4; cat kept in a block of flats, odds ratio 3.6, 95% CI 1.2 to 10.8) (Tenca 2016). Our own U.S. data agree with that null: percent rural population, private well water and cumulative pesticide application show no association (Figure 5). We therefore do not rest the hypothesis on rural residence. The claim we make is narrower and stronger, namely that the association is specific to Ixodes rather than to rural or outdoor exposure generally. Pet contact is not inconsistent with this, since companion animals carry ticks into the household; our canine Anaplasma seroprevalence replicate is a direct measure of that route.

In North America and Europe, the regions with the highest reported PSC incidence seemed to overlap with regions endemic for *Ixodes* ticks. We therefore set out to confirm this suspicion.

This pattern suggests the hypothesis that PSC is tied to exposure to a pathogen or antigen transmitted by *Ixodes* ticks. In theory, this exposure could trigger an inflammatory and/or autoimmune process targeting the biliary and/or colonic epithelium. Such a process might then be perpetuated by some other driver within the environment, gut or immune system. Genetic susceptibility can predispose to this process but is neither necessary nor sufficient for disease. We examine several major alternative explanations (latitude/vitamin D, enteric and waterborne pathogens, pesticides, air pollution, rural residence, and healthcare-access artifacts).

Finally, we present several concrete, testable predictions made by the hypothesis.

## Methods

We examined several independent ecological datasets. First, a between-country comparison of population-based PSC incidence versus tick-exposure markers was done. Second, we performed a within-country U.S. state-level analysis derived from individual-level insurance claims, as well as within-country data from Italy and Japan. Third, we reviewed veterinary surveillance data for independent corroboration.

### Between-country data

#### PSC, AIH, and PBC incidence

Incidence estimates were drawn from peer-reviewed, population-based studies that defined a geographic catchment, used explicit diagnostic criteria^1^, and had data sufficient to calculate an annualized rate. We prioritized more recent, population-based designs and avoided overlapping catchments; the primary analysis uses one estimate per region. Fourteen population-based incidence estimates across North America, Europe, and Oceania were compiled. Thirteen of these regions have comparable population-based Lyme-incidence data, and eleven of those have established *Ixodes* populations; those eleven form the primary between-country PSC–Lyme comparison.

#### *Ixodes* exposure

Standardized tick-density data across countries are sparse, so we used Lyme disease incidence as a proxy for human exposure to *Ixodes*-endemic environments. It is unlikely that PSC is triggered by *Borrelia* (see Discussion) but its transmission stands as a proxy for that of numerous other tick-borne microbes. Where possible, we relied on laboratory-confirmed surveillance (for example, disseminated Lyme borreliosis or neuroborreliosis), which is more specific than erythema migrans diagnoses. For several countries without national laboratory-confirmed Lyme surveillance we used the best available reported incidence.

#### HLA-DRB1*03 frequency and latitude

HLA-DRB1*03 (DR3) allele frequencies were compiled from the Allele Frequency Net Database and population-based studies^12^; latitude was the catchment centroid.

### Within-country data

#### United States

PSC, AIH and PBC rates were obtained from the Merative MarketScan Commercial Claims and Encounters (CCAE) and Medicare Supplemental (MDCR) databases for 2018–2022. The study window begins on 1 October 2018, when the PSC-specific International Classification of Diseases, 10th Revision, Clinical Modification (ICD-10-CM) code K83.01 was introduced. Prevalent cases required ≥ 2 distinct service dates carrying K83.01 ≥30 days apart within a 12 month period. Incident cases additionally required the first-ever K83.01 claim within the index year and ≥12 months of prior continuous enrollment. AIH and PBC were identified by equivalent criteria. Over 110.7 million person-years we identified 5,531 prevalent / 1,135 incident PSC cases, 19,548 / 2,347 AIH cases, and 13,083 / 1,590 PBC cases.

State- and national-level rates were age- and sex-standardized using the pooled MarketScan enrollee distribution. Each state’s five-year (2018–2022) pooled counts were used; if the total was fewer than 11 cases, the count was not used in the correlation analyses. State-level data for all three diseases are given in Supplementary Table S3.

#### Italy

Regional PSC incidence (2012–2014) is from the Italian National Rare Diseases Registry.^13^ Regional *Ixodes* density was classified on a 7-point ordinal scale from ECDC distribution maps and published tick surveys; it is the primary within-Italy exposure measure. As a complementary check, we used regional Lyme case rates from a multi-center series of 1,260 Lyme borreliosis cases from eight hospital centers across five regions (2010–2022), a center-ascertained case-rate proxy, not population-based incidence.^14^

#### Japan

Prefecture-level PSC prevalence was provided by A. Tanaka (Intractable Hepatobiliary Disease Study Group, Teikyo University School of Medicine, Tokyo; personal communication, January 2026), derived from Japan’s national medical expense subsidy program. The Japanese analysis is prevalence-based. The subsidy certifies only patients whose disease is severe enough to qualify, so mild PSC is not captured. Prefecture-level *Ixodes* and *Haemaphysalis longicornis* densities were assigned on a 0–3 ordinal scale from published Japanese tick-distribution surveys, and Lyme disease incidence was taken from national surveillance data.^15^

#### Environmental exposures

Twenty-four state-level exposures (Supplementary Table S4) were constructed from public data sources, spanning *Ixodes*-borne pathogen incidence, tick habitat, non-*Ixodes* tick-borne pathogens, tick ecology, latitude, smoking, agricultural land, water, air-pollution, and proximity of liver specialists and transplant centers. Full source details and construction are given in Supplementary Note S1.

#### Statistical analysis

For each exposure we computed Spearman and Pearson correlations with PSC rates. Our one a priori adjustment in the U.S. was for European ancestry; we singled this variable out because it is a non-tick factor that both varies geographically and independently raises PSC risk through the HLA-B*08:01–DRB1*03:01 haplotype. We estimated each state’s European-ancestry fraction from the American Community Survey and held it constant when correlating exposures with PSC. We use ancestry rather than HLA directly because no measured state-level HLA data exist.

Between countries, where n = 11 and the Lyme values are heterogeneous in provenance, only the ranking is interpretable, so the Spearman rank correlation is the primary measure. The United States analysis used a Pearson partial correlation adjusted for European ancestry, computed on log10(x + 0.1) for the count-based surfaces (tick-borne disease incidence and cumulative pesticide application), untransformed for the remaining continuous surfaces. The two veterinary correlations were unadjusted.

We deliberately did not adjust for census region in the main analysis: since the hypothesis is that PSC clusters geographically *because* tick exposure does, controlling for region would remove the very signal we are testing. For *Anaplasma* we ran an additional test that also held tick density constant, asking whether where people are actually being infected predicts PSC over and above where tick habitat merely exists.

We tested whether PSC clustered geographically using Moran’s I, a standard measure of spatial correlation. Spatial weights are queen-contiguity and row-standardized. Because states with counts <11 are suppressed, omitting them can leave a state with no neighbor for reasons of data availability rather than geography. Maine is the only such state, and analyses were done with and without it.

All analyses were run in MATLAB. All research was conducted in accordance with the Declarations of Helsinki and Istanbul. The MarketScan analysis used fully de-identified secondary data and was determined to be exempt by the Yale University IRB #2000043343.

## Results

### Cross-country: PSC incidence correlates with Lyme disease incidence

Across *Ixodes*-endemic regions of Europe and North America, PSC incidence shows a geographic concordance with reported Lyme disease incidence (Figure 1). The regions reporting the highest PSC incidence worldwide, Finland (1.58 per 100,000), Sweden (1.8), and Olmsted County, Minnesota (1.47), are also regions with long-established, high-density *Ixodes* populations and substantial Lyme burden; the lowest PSC rates occur in Mediterranean Europe, where Lyme is rare.

**Figure 1.**
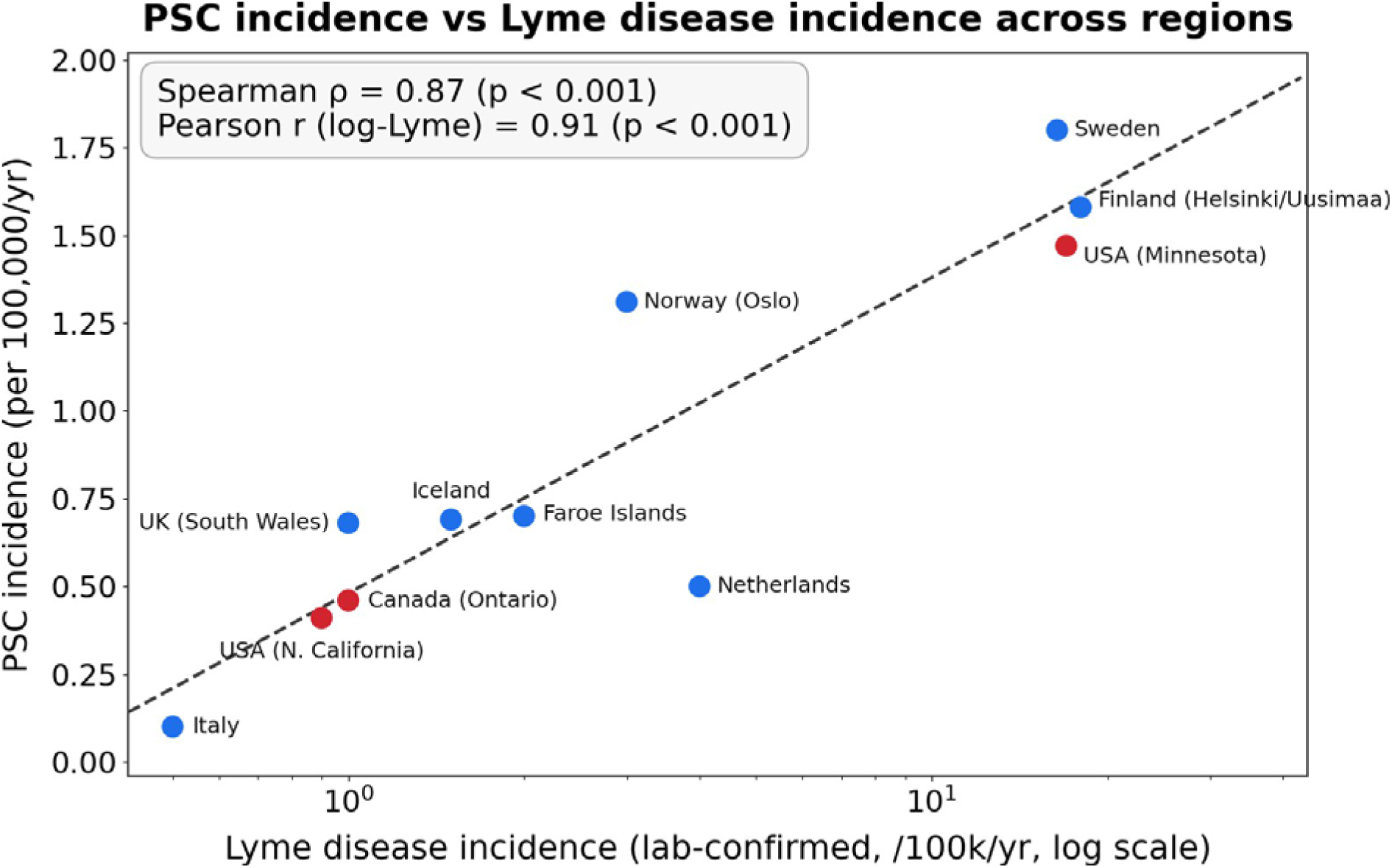
Cross-country PSC incidence versus Lyme disease incidence (log scale). Lyme is used only as an available proxy for Ixodes tick exposure here; tick density would be preferable but is not generally available across European data sources. Among the 11 regions, Spearman ρ = 0.87 (Pearson on log-Lyme r= 0.91). PSC does occur where classical Lyme is rare (e.g. Australia, New Zealand) but where various pathogen candidates are shared across certain tick species (see text).

The Spearman correlation between PSC and Lyme incidence is ρ = 0.87 (p < 0.001), and the Pearson correlation on log-transformed Lyme incidence is r = 0.91 (p < 0.001; Table 1).

**Table 1.** Regional PSC, AIH, and PBC incidence with reported Lyme incidence, HLA-DRB1*03 frequency, and latitude.

| Region | PSC | AIH | PBC | Lyme | HLA-DR3 % | Latitude |
|---|---|---|---|---|---|---|
| Finland<sup>6,16</sup> | 1.58 | 1.1 | 1.70 | 18 | 9.7 | 60°N |
| Sweden<sup>17,18</sup> | 1.80 | 0.85 | 2.30 | 16.4 | 13.1 | 62°N |
| Norway<sup>19</sup> | 1.31 | 1.90 | 1.60 | 3 | 13.5 | 60°N |
| Faroe Islands<sup>20</sup> | 0.70 | 5.2 | 2.5 | 2 | 10.2 | 62°N |
| Iceland<sup>21–23</sup> | 0.69 | 2.20 | 2.00 | 1.5‡ | — | 64°N |
| UK<sup>24–26</sup> | 0.68 | 1.94 | 2.2 | 1.0 | 14.8 | 54°N |
| Netherlands<sup>27,28</sup> | 0.50 | 1.1 | 1.10 | 4 | 13.2 | 52°N |
| Italy<sup>13,29</sup> | 0.10 | — | 5.31 | 0.5 | 9.95 | 43°N |
| Canada–Ontario<sup>30</sup> | 0.46<sup>§</sup> | — | — | 1.0 | — | 44°N |
| USA–Minnesota<sup>7,31</sup> | 1.47 | — | 2.70 | 17 | 13.7 | 45°N |
| USA–N. California<sup>8</sup> | 0.41 | — | 2.75 | 0.9 | 14.5 | 39°N |
| New Zealand<sup>32–34</sup> | 0.77 | 1.90 | 0.8 | ‡ | — | 41°S |
| Greenland<sup>10</sup> | 0 | present | present | ‡ | — | 64°N |
| USA–Alaska Natives<sup>9</sup> | 0 | 42.9* | 16.0* | ‡ | 0.6 | 61°N |

\* prevalence per 100,000 (excluded from incidence-based correlations)
§ PSC-IBD incidence, not all PSC cases.
‡ Lyme is rare and travel-associated.
See Supplement Note S4 for further detail. Superscripts refer to the corresponding references.

### PBC and AIH do not correlate with Lyme disease incidence

The PSC–Lyme correlation is specific to PSC; neither AIH (ρ = −0.67, p = 0.07) nor PBC (ρ = −0.11, p = 0.75) tracks Lyme incidence (Figure 2), so it is not a generic autoimmune-liver-disease signal.

**Figure 2.**
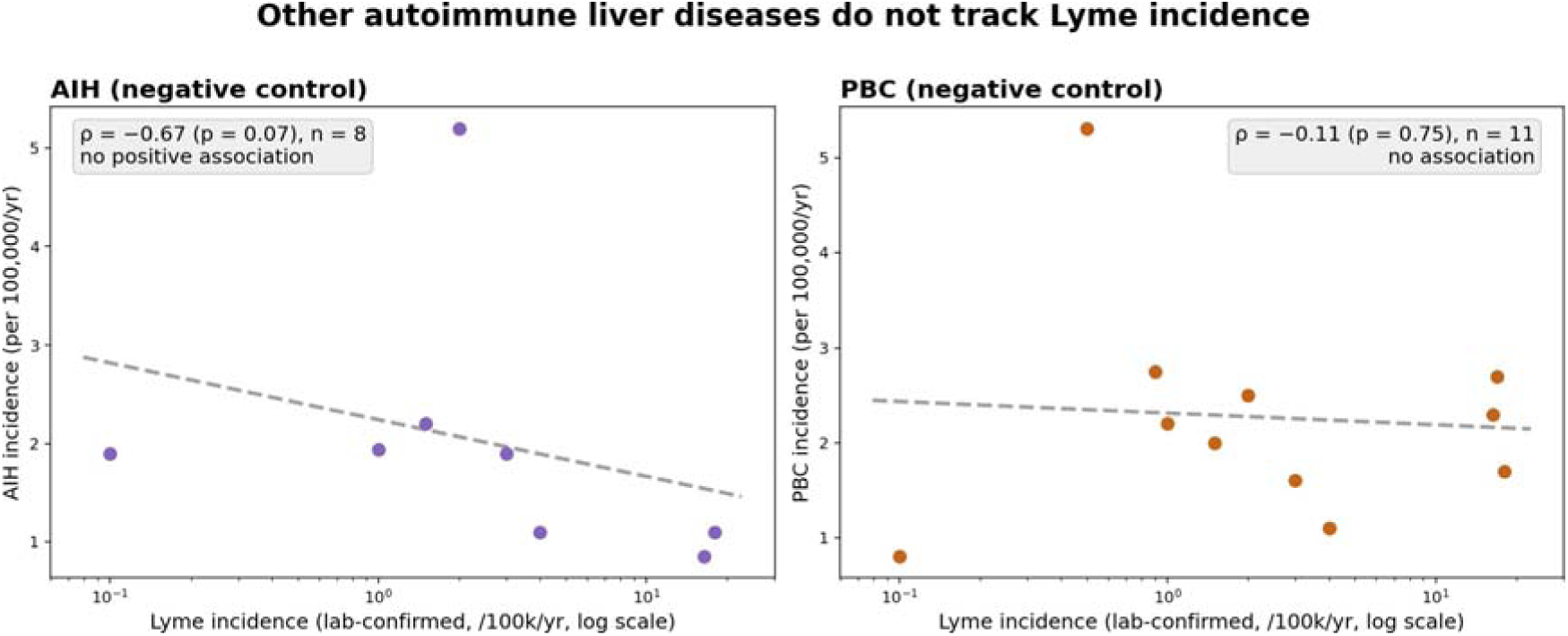
Neither autoimmune hepatitis (AIH, ρ = −0.67, p = 0.07, n = 8) nor primary biliary cholangitis (PBC, ρ = −0.11, p = 0.75, n = 11) tracks reported Lyme incidence across regions, indicating that the PSC–Lyme association is not a generic autoimmune-liver-disease signal. Each point is one region from Table 1. Alaska is excluded from both controls because its AIH and PBC values are prevalence proxies rather than incidence.

### HLA association

In this cross-country analysis, population HLA-DRB1*03 frequency did not track PSC incidence across regions (ρ = 0.13, p = 0.73; Pearson r = 0.44, p = 0.21). This is not evidence against the HLA association, which is among the best-replicated in hepatology at the individual level. It reflects the limits of an ecological comparison: DRB1*03 frequency varies only 1.5-fold across the European-descended populations in Table 1 (9.7–14.8%) against a 36-fold range in reported Lyme incidence, and with n = 10 the design can detect only |r| > 0.63. We therefore adjusted for European ancestry directly in the U.S. analysis (Methods) rather than relying on this null.

Direct evidence that ancestry rather than place accounts for part of the geographic variation comes from New Zealand: within the single Canterbury region, PSC prevalence was 14.35 per 100,000 in people of European ethnicity versus 5.45 per 100,000 in non-Europeans.32 A 2.6-fold gap within one geography, one health system and one surveillance method is the clearest available justification for adjusting for European ancestry, and all U.S. associations reported below are ancestry-adjusted.

### Alaska and Greenland: No Ixodes pathogens, No PSC

Alaska Native and Greenlandic populations report essentially zero PSC, despite the fact that they share many other features with high-PSC populations: cold climate, similar diets, livestock exposure (reindeer and caribou herding, sheep), rodent exposure, and high rates of waterborne illness.^37^ Neither has established transmission of *Ixodes*-borne pathogens to humans; Alaska’s established ticks (chiefly *Ixodes angustus*) are primarily small-mammal-associated and are not established vectors of Lyme disease or anaplasmosis. AIH incidence in Alaska Natives is among the highest in the world, at 42.9–58.6 per 100,000,^9^ and Greenland reports both AIH and PBC.^10^ Both have IBD at rates comparable to other northern populations. These populations show that autoimmune-liver-disease susceptibility, IBD, and multiple alternative environmental exposures can all be present while PSC is absent. A distinguishing feature is the absence of established human exposure to Ixodes-borne pathogens (Supplementary Table S2).

Ascertainment deserves explicit comment, because PSC is diagnosed cholangiographically whereas AIH and PBC are diagnosed serologically and histologically. The autoimmune liver disease diagnoses in both reports were histologically confirmed, but neither population has undergone systematic magnetic resonance cholangiography screening of its IBD cohort.

Screening studies elsewhere identify PSC in 8.1 to 9.0% of adults and 15.1% of children with IBD, against 0.6 to 4.3% by clinical ascertainment (Lunder 2016; Bowlus 2023), so an unscreened zero is a weaker observation than a screened one. Magnetic resonance cholangiography screening of the Alaska Native or Greenlandic IBD population would convert this argument into a test, and we list it among the predictions below.

### Latitude and Vitamin D Deficiency

PSC’s relationship to latitude is weaker than it first appears, and the best-controlled analysis argues against a latitude mechanism altogether. In 8,590,421 UK primary-care records, PSC incidence did not vary across 50°N to 57°N (0.82 to 1.02 per 100,000 per year, p = 0.473; adjusted r2 = 0.053, p = 0.582), whereas PBC (2.16 to 4.86, p = 0.002) and AIH (2.00 to 3.28, p = 0.003) both rose with latitude in the same cohort.25 This internally controlled dissociation mirrors our own control analysis: the two diseases that do track latitude are the two that do not track Lyme incidence, and PSC behaves in the opposite way on both axes. Alaska and Greenland are further exceptions, as noted. Finland lies at a high latitude and reports one of the highest PSC incidences worldwide6. At the same time however, tick densities and tick-pathogen carriage are among the highest in Europe, with both Ixodes ricinus in the south and Ixodes persulcatus in the north.38-39 Finland has extensive outdoor recreation: the country’s ‘Everyman’s Right’ grants universal forest access, with over 55-60% of Finns participating in berry picking and mushroom foraging during peak tick activity season.40 Finland also has high HLA-B*08/DR3 frequency (22-25%).

The latitude pattern raises the possibility that vitamin D deficiency could underlie the observed disease incidence. However, a paradoxical north-south vitamin D gradient exists in Europe: Northern European populations show superior vitamin D status compared to Southern Europe due to cod liver oil consumption, food fortification, and fatty fish intake.^41^ Finland, with one of the highest PSC incidence rates globally, also has among the best vitamin D status in Europe.^42^ Conversely, the Eastern Mediterranean region has the highest global prevalence of vitamin D deficiency yet low PSC rates.^43^ Additional latitude-associated factors including ultraviolet radiation exposure, photoperiod effects, and various infectious agents may covary with latitude but their contributions remain speculative.

### United States: PSC prevalence is geographically clustered

In the United States, PSC prevalence is not geographically random. Three high-prevalence clusters are evident (Figure 3A): a Northeastern through upper-Midwest cluster, a Northwestern cluster, and an intermountain cluster (Utah, Colorado). The clustering is significant on raw prevalence (global Moran’s I = 0.39, permutation p = 0.002) and remains so after European ancestry is accounted for (residual Moran’s I = 0.40, permutation p = 0.001). Across the 36 states, five-year-pooled, age- and sex-standardized prevalence varied roughly four-fold, from 2.2 per 100,000 in Oklahoma to 9.0 in Maine; the national standardized prevalence was 5.72 per 100,000 in 2022 and incidence 0.42 per 100,000, both likely underestimates (see Limitations). States with fewer than 11 five-year-pooled cases (gray) are excluded from correlations.

**Figure 3.**
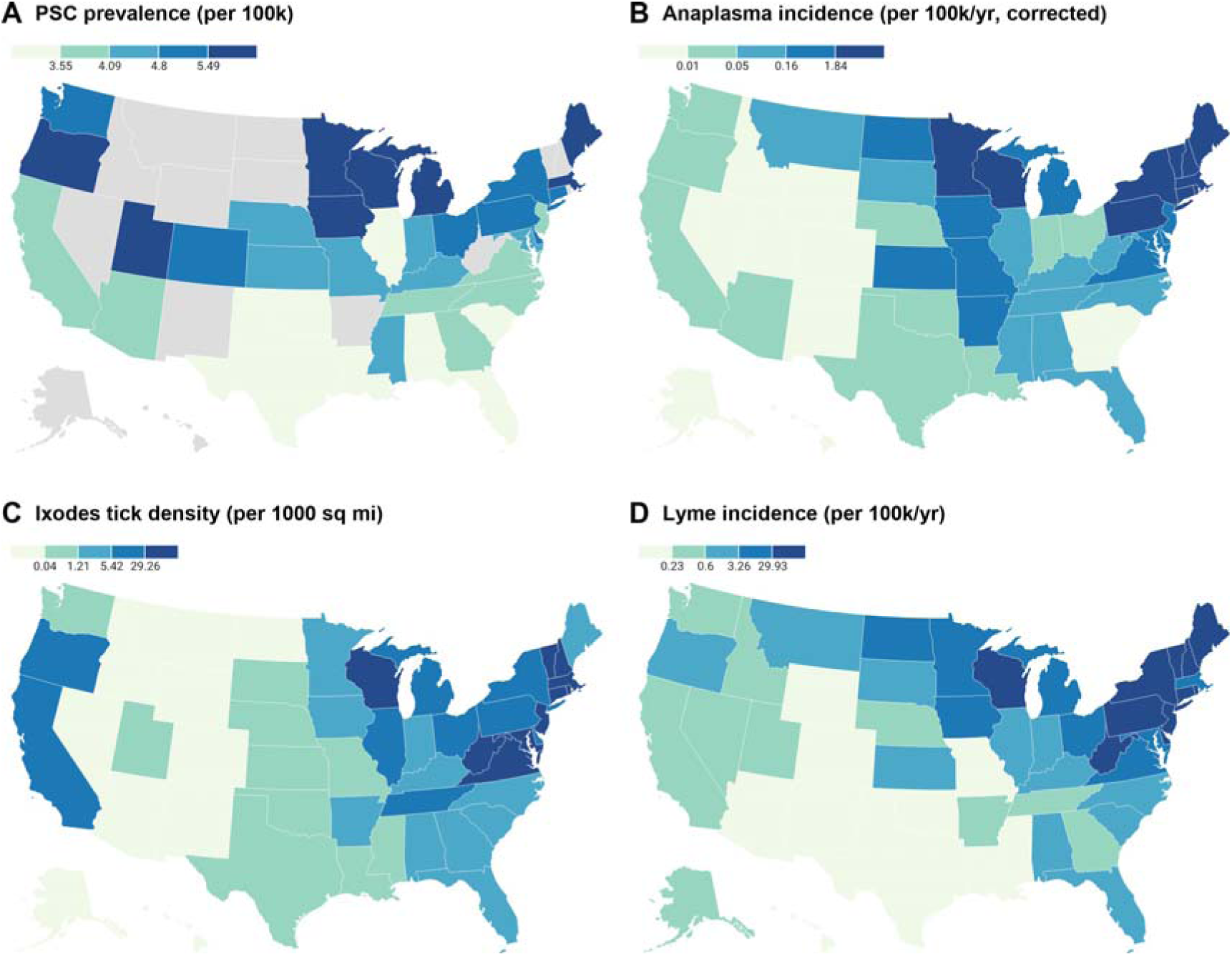
The geographic distribution of PSC in the United States (A, prevalence) parallels *Ixodes*-borne *Anaplasma* incidence (B), *Ixodes* tick habitat (C), and Lyme incidence (D). Colors represent quintiles of each panel’s distribution. Gray states in (A) have fewer than 11 PSC cases, suppressed under the MarketScan data-use agreement. Sources: MarketScan 2018–2022 (PSC); CDC surveillance (*Anaplasma*, Lyme); GBIF (tick density).

### Correlation with tick habitat

PSC prevalence rises with *Ixodes* habitat as mapped by the Global Biodiversity Information Facility (GBIF), though this falls just short of significance on the primary log scale (ancestry-adjusted partial r = +0.26, 95% CI −0.08 to +0.55); the CDC reference standard for established *I. scapularis* or *Ixodes pacificus* ³ gives a stronger, significant correlation (+0.48, p = 0.003), and the two habitat sources agree closely (Spearman ρ = 0.84).

### Correlation with tick pathogen incidence

PSC prevalence tracks *Ixodes*-borne pathogen transmission, not tick habitat alone (Figure 3B, D). Human anaplasmosis (*Anaplasma phagocytophilum*) incidence correlates with PSC prevalence at an ancestry-adjusted partial r = +0.50 on the log scale (p = 0.002; Spearman ρ = +0.33) (Figure 4). Holding *Ixodes* density constant leaves the correlation essentially unchanged (partial r = +0.49). In a joint regression the anaplasmosis coefficient (+0.61) exceeds that of *Ixodes* density (+0.36). All three *Ixodes*-borne pathogens converge on the same magnitude (*Anaplasma* +0.50, *Babesia* +0.56, Powassan virus +0.52), whereas every non-*Ixodes* tick-borne infection is negative (Supplementary Table S4). Within the uniformly high-latitude Northeast– Midwest block, *Babesia* and Powassan virus behave likewise (r = +0.84 and +0.78, ancestry- and latitude-adjusted; Spearman ρ = +0.55 and +0.50, p = 0.05 and 0.07 respectively). The correlation with Lyme incidence is comparable, again just short of significance on the log scale (ancestry-partial r = +0.33; 95% CI −0.01 to +0.59).

**Figure 4.**
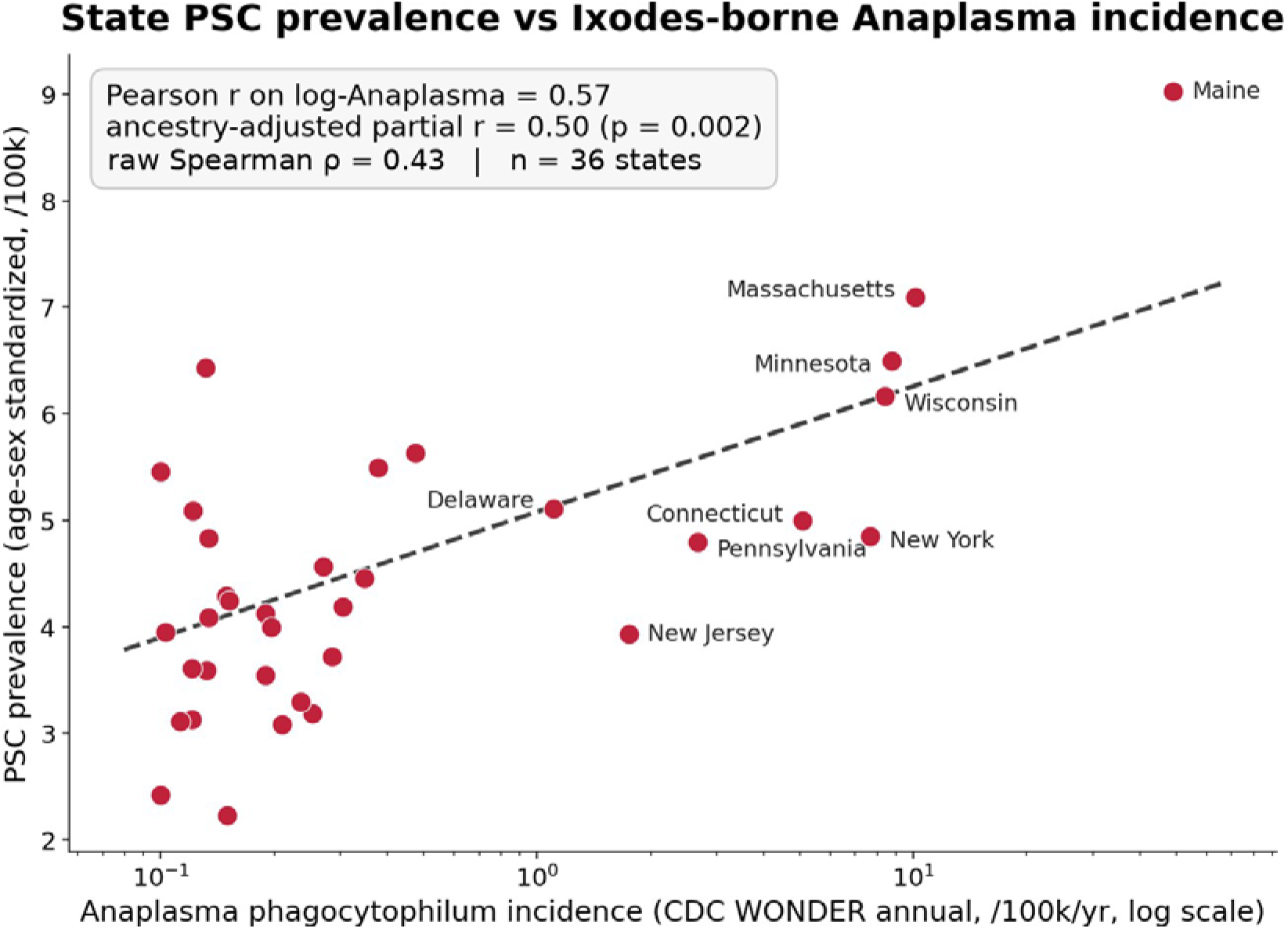
State PSC prevalence versus *Ixodes*-borne *Anaplasma phagocytophilum* incidence (CDC WONDER annual, 2018–2021). Pearson r on log-*Anaplasma* = 0.57; ancestry-adjusted partial r = 0.50 (p = 0.002); raw Spearman ρ = 0.43 (ancestry-adjusted partial ρ = 0.33); n = 36 states.

**Figure 5.**
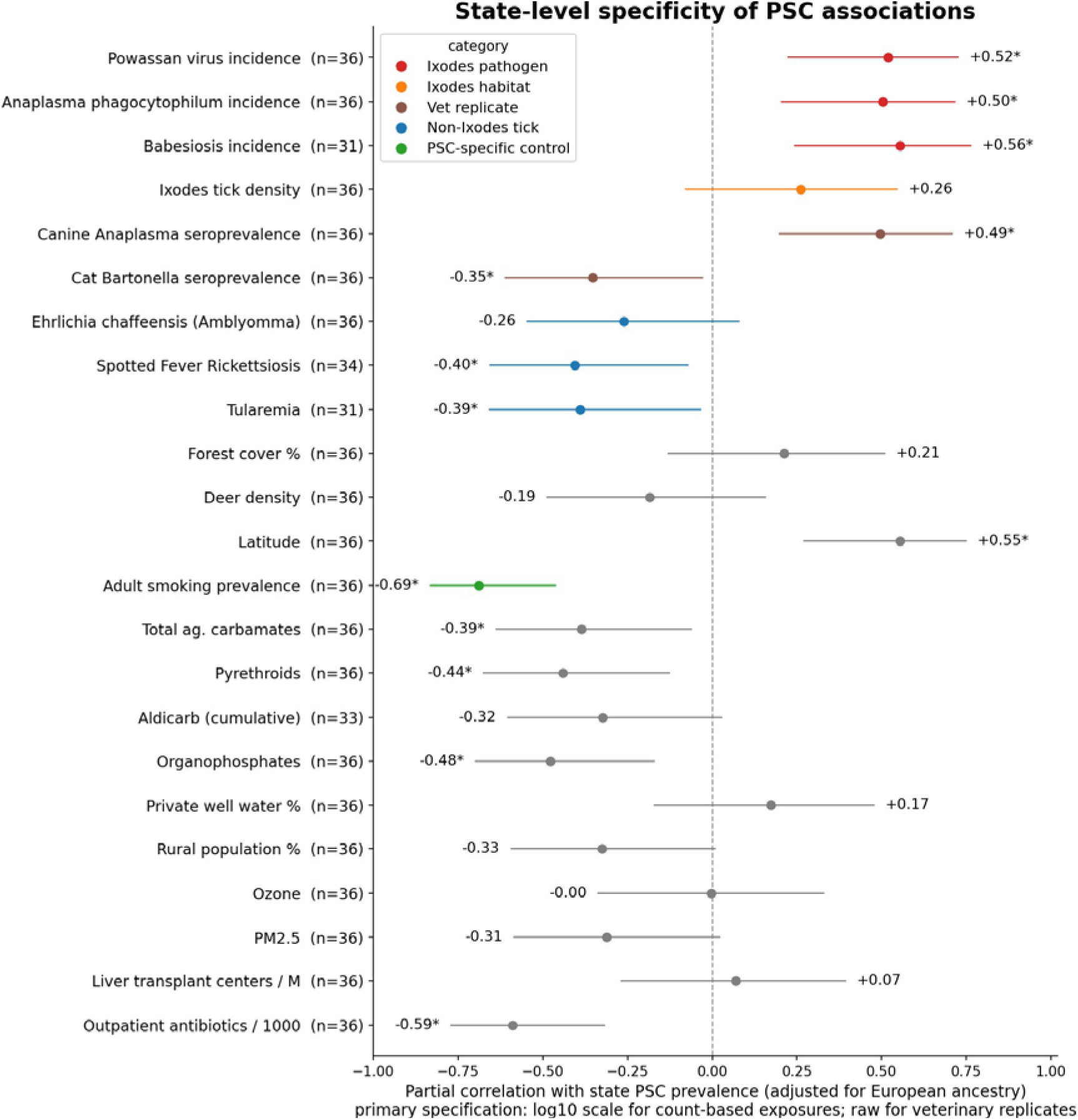
State-level specificity of PSC associations, adjusted for European-ancestry fraction. *Ixodes*-borne pathogens are positively associated with PSC prevalence, whereas non-*Ixodes* tick-borne infections and the autoimmune-liver and environmental controls are null-to-negative.

The intermountain cluster is the clearest counterexample within the U.S. data. Utah and Colorado carry high PSC prevalence yet report little anaplasmosis and few *Ixodes*. Two explanations are plausible but unmeasured: Utah has among the highest European-ancestry fractions in the country, which our linear ecological adjustment may not fully remove; and a large proportion of its population spends up to two years on missionary work outside the state, so tick-borne exposure may be real but invisible to in-state surveillance. The discriminating test is serologic rather than ecological: if intermountain PSC patients show no more evidence of *Ixodes*-borne infection than local controls, the cluster is a genuine exception and the hypothesis is narrower than we propose; if they do, the exposure was simply unmeasured.

### Veterinary replication of anaplasmosis association with PSC

In the United States, canine *Anaplasma phagocytophilum* seroprevalence^44^ (IDEXX, ∼60-million-test database) correlates with human PSC prevalence (raw r = +0.49, weighted +0.59, p = 0.002). It also agrees with the human *Anaplasma* signal. Conversely, cat *Bartonella henselae* seroprevalence^45^ is *inversely* correlated with PSC (r = −0.35, p = 0.036). *B. henselae* exposure concentrates in the South and is low in the Northeast and Pacific, the opposite of PSC. This argues against *Bartonella* or cat-scratch exposure as a population-scale PSC trigger (Supplementary Table S4).

### Specificity of the *Ixodes* association

Multiple environmental exposures are shown in Figure 5 (United States; see Supplementary Table S4 and Supplementary Note S2).

*Ixodes*-borne pathogens (*Anaplasma* +0.50, *Babesia* +0.56, Powassan virus +0.52) are positively associated with PSC prevalence. Non-*Ixodes* tick-borne infections do not behave like the *Ixodes*-borne ones. *Ehrlichia chaffeensis* (transmitted by *Amblyomma americanum*) is null (ancestry-partial r = −0.26, not significant), and spotted fever rickettsiosis (−0.40) and tularemia (−0.39; both *Dermacentor*/*Amblyomma*-associated) are inversely related to PSC. A confounder operating through wooded, outdoor, or rural exposure would not distinguish *Ixodes*-transmitted from *Amblyomma*- or *Dermacentor*-transmitted infections.

AIH and PBC prevalence are uncorrelated with *Ixodes* density, and their incidence is null-to-negative against Lyme. Their per-state geography differs from PSC’s (AIH highest in Tennessee, Michigan, Utah, Alabama, Wisconsin; PBC highest in Nebraska, Kansas, Texas, Indiana, Georgia). This argues against a shared autoimmune or healthcare-access confounder.

PSC is known to be *inversely* associated with smoking at the individual level, and the MarketScan data reproduces that known signature (ancestry-adjusted partial r = −0.69, p < 0.001). The smoking correlation is null for AIH (+0.08) and PBC (−0.04). Recovering an established, disease-specific association is reassuring about the MarketScan data quality.

Points are ancestry-adjusted partial correlations with 95% confidence intervals on the primary specification (log10 scale for count-based exposures; raw and unadjusted for the two veterinary replicates); asterisks mark p < 0.05.

### Within-country analyses: Italy and Japan

In Italy, regional PSC incidence is higher in the north and falls toward the south, tracking *Ixodes density* (ρ = 0.68) across the 19 regions with a PSC estimate (Figure 6). A Lyme case-rate proxy^14^ shows the same gradient (ρ = 0.61, p = 0.004). We exclude Puglia’s 2013 year count of 24 PSC cases versus 1 case in 2012 and 6 in 2014; because the Italian National Rare Diseases Registry was still being populated over this period, we read the spike as a batch upload of previously diagnosed cases. The gradient is insensitive to this choice: for *Ixodes* density ρ = 0.64 with Puglia’s 2013 count retained, 0.68 with the 2013 count removed. The Lyme-proxy gradient is 0.55, 0.61, and 0.62 respectively (all p ≤ 0.012, not shown).

**Figure 6.**
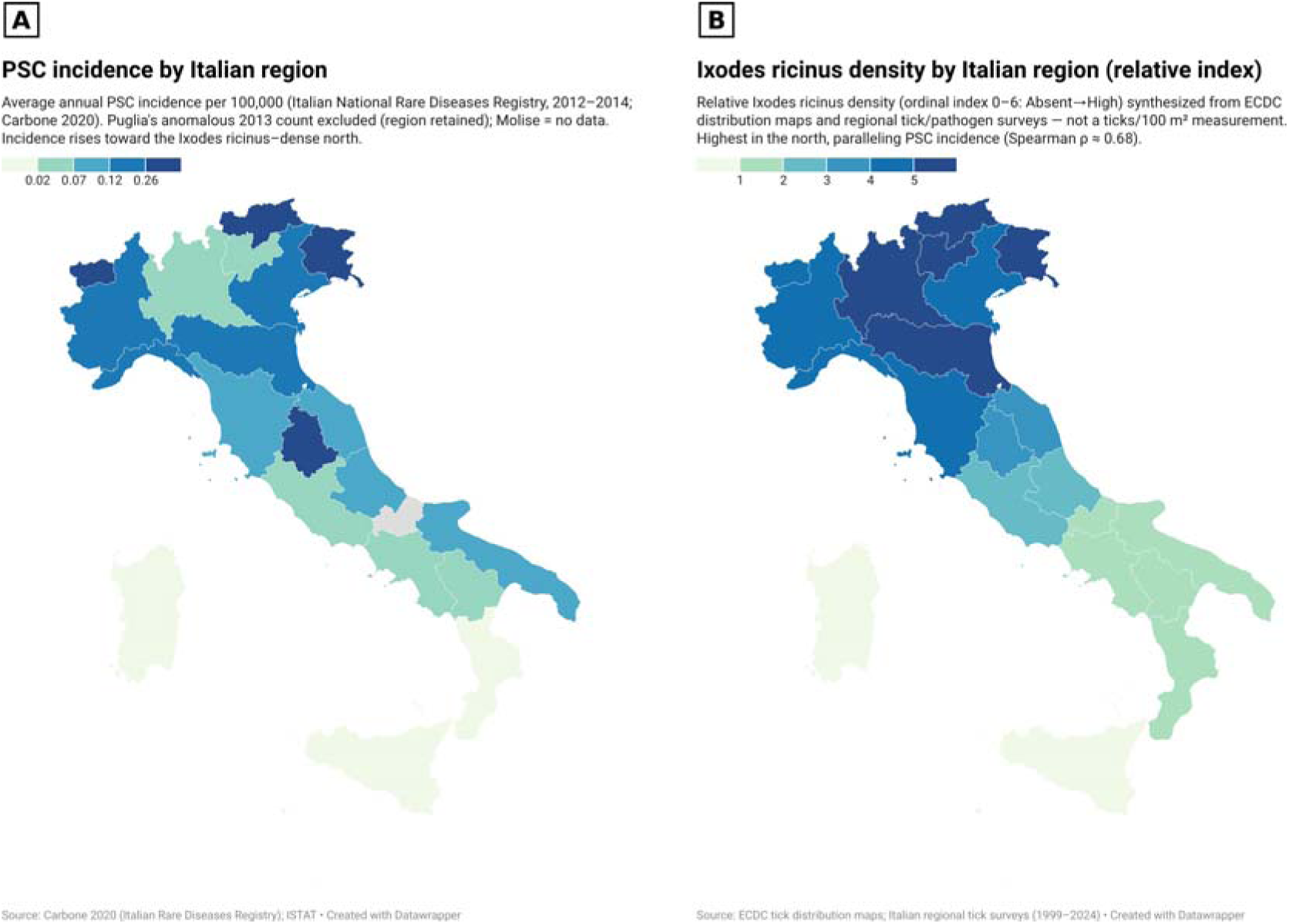
Italy, within-country gradient. Regional PSC incidence (A) tracks *Ixodes* density (ρ = 0.68) (B) and a clinical-series Lyme case-rate proxy (ρ = 0.61 not shown) across 19 regions (Molise no data; Trentino-Alto Adige split into its two provinces, n = 20). Puglia’s 2013 registry backfill was removed; the gradient is robust to its inclusion (see text). Panel A is shaded in quintiles; Panel B shows the ordinal *Ixodes density* class (absent→high). See Supplementary Table S5. Maps built with Datawrapper.

In contrast, in Japan the data do not support the hypothesis. Prefecture-level PSC prevalence is uncorrelated with *Ixodes* density (ρ = −0.02), *Haemaphysalis longicornis* density (ρ = −0.15), or Lyme incidence (ρ = −0.03). The two tick measures combined are also uncorrelated (ρ = −0.13; Supplementary Table S6).

A related caution comes from China, where PSC period prevalence of 2.36 per 100,000 shows a five-fold internal gradient (East China 4.87 versus South China 0.92) that tracks per-capita gross domestic product (rate ratio 1.65) and healthcare expenditure (1.94) rather than any ecological variable (Xu 2024). Economic and diagnostic-access gradients can therefore generate within-country PSC variation on their own. Our U.S. design addresses this directly: adult liver-transplant-centre density and outpatient antibiotic prescribing, both proxies for healthcare access and case-finding intensity, show no association with PSC prevalence (Figure 5).

We report this Japanese null as a genuine challenge to the hypothesis rather than an artifact to be explained away. If the finding holds up to further investigation it must be held as a major blow against the hypothesis. Several alternative explanations merit consideration however: (1) the relationship may be geographically restricted to North America and Europe, whose human-biting *Ixodes* species and pathogen assemblages differ from Japan’s; (2) the available Japanese exposures may be inadequate proxies; notably, anaplasmosis, one of the stronger U.S. correlates, is not systematically surveilled in Japan; (3) a long exposure-to-disease latency could decouple current prevalence from current tick surveillance; (4) the prevalence measure derives from a national subsidy that records only more severe disease, and prefecture case counts are small.

### Oceania

Australia and New Zealand have PSC but no endemic Lyme disease. They do carry *Ixodes* species; the endemic *Ixodes holocyclus* carries *Candidatus Neoehrlichia australis* and Ca. N. *arcana*, *Anaplasmataceae* congeneric with the European pathogen Ca. N. *mikurensis*.^46^ Both countries also have *Haemaphysalis longicornis* (family *Ixodidae*), which harbors pathogens overlapping those of *Ixodes*, including *Anaplasma phagocytophilum*. However, human transmission and pathogenicity of these Australian organisms are undetermined. We therefore treat Oceania not as supporting evidence but as a prediction: if these organisms prove human-transmitted and pathogenic, PSC-endemic Oceania would fit the hypothesis, and demonstrating their pathogenicity, or its absence, is a direct test.

## Discussion

### Summary and interpretation

We present evidence for a testable hypothesis that PSC is connected to tick exposure, specifically to *Ixodes* ticks and to functionally analogous hard-tick vectors that transmit the same group of pathogens.

Across regions in Europe and North America, PSC incidence correlates with Lyme disease, a proxy for *Ixodes* tick exposure, whereas AIH and PBC incidence do not. Within the United States, PSC prevalence correlates with tick habitat and *Ixodes*-borne pathogen incidence (*Anaplasma*, *Babesia*, Powassan virus), whereas non-*Ixodes* tick-borne infections do not.

Latitude co-varies with *Ixodes* endemicity but is dissociated from PSC by Alaska and Greenland, which show abundant non-PSC liver autoimmunity but no established human exposure to *Ixodes*-borne pathogens and no PSC. No correlation was seen with numerous other environmental exposures. Analyses were adjusted for European ancestry as a proxy for the PSC-associated HLA haplotypes.

No generic “rural,” “wooded,” or “Northern” confounder predicts the constellation of our findings. Multiple comparisons can produce an isolated false positive; they less readily produce a coherent, directional pattern. Correlated exposures and residual spatial confounding remain possible explanations we cannot fully exclude.

### Candidate triggering pathogens

Ticks carry a large array of microbes^47^, many of which remain of unclear character and clinical significance. More than one pathogen might contribute to disease; co-infection of ticks is common^48^ and co-infection of humans is increasingly being recognized. Different geographic regions may harbor different but functionally analogous agents that share a common pathogenic trait, such as a critical antigenic feature. For example, lipopolysaccharide (LPS) cross-reactivity among intracellular bacteria is well documented.

Here we focus on candidate triggers (Table 2) that are (i) carried by *Ixodes* or a functionally analogous hard tick, defined as a human-biting *Ixodidae*, (ii) capable of biliary or hepatic involvement or of immune cross-reactivity with biliary antigens, and (iii) compatible with the geographic and specificity pattern above.

**Table 2.** Candidate tick-borne triggering pathogens with documented biliary or hepatic involvement (references in text)

| Pathogen | Tick vectors | Tropism | Evidence for biliary / hepatic involvement | Prevalence in ticks |
|---|---|---|---|---|
| *Anaplasma phagocytophilum* | *I. ricinus*, *I. scapularis*, *I. persulcatus* | Granulocytes; periportal | Periportal lymphohistiocytic infiltrates; abnormal LFTs ~67% | 1–25% (Scandinavia); ~20% Europe (ticks / reservoirs) |
| Ca. *Neoehrlichia mikurensis* / *australis* | *I. ricinus*; *I. holocyclus* | Vascular endothelium | Endothelial culture; vasculitis / thrombosis; FtsZ mimicry of β-tubulin (p-ANCA target) | Up to 25% |
| *Chlamydia*-like organisms | *I. ricinus* | Intracellular; endothelial | Anti-*Chlamydia*-LPS IgA in PSC (≈80% positive; OR 6.7, 95% CI 3.0–17.0); tick-to-human transmission shown | 40% adult *I. ricinus* (Finland); untested in *I. scapularis* |
| *Bartonella henselae* | *I. ricinus* (and cats) | Endothelial; biliary | Lymphocytic cholangitis in cats; BUT inverse state-level correlation argues against population-scale role | Present *I. ricinus*; absent Scandinavian ticks |
| *Borrelia burgdorferi* | *I. scapularis*, *I. ricinus*, *I. persulcatus* | Disseminated | Excluded as sole trigger: PSC occurs where Lyme is absent; serves as proxy for *Ixodes*-endemic residence | 10–30% *I. ricinus*; 15–25% *I. scapularis* |
| Tick saliva / alpha-gal antigens | *Ixodes* spp., *Haemaphysalis* spp. | Immune sensitization | Tick bites induce durable IgE to galactose-α-1,3-galactose; alpha-gal IgE proposed as exposure marker | hard-tick saliva |

*Anaplasma*, *Babesia* and Powassan virus have nearly identical correlations with PSC (+0.50, +0.56, +0.52), so the data point to the vector rather than any one pathogen. A currently unknown agent could well be responsible. A role for tick saliva cannot be excluded. These candidates are discussed further in Supplementary Note S5.

**Tick saliva.** If a tick-induced antigen matters, saliva is as plausible a source as a pathogen. Hard-tick bites can induce durable immune sensitization. The tick salivary proteome contains Salp15, sialostatins, glycine-rich cement proteins, histamine-binding proteins and more. Both *Ixodes* and *Haemaphysalis* saliva cause alpha-gal syndrome linked to carbohydrate antigens. Measuring alpha-gal IgE in PSC cohorts would give a marker of tick-saliva exposure.

We suggest an etiologic model in the form of a “trigger-driver” mechanism (Supplementary Figure S1). A tick bite delivers a pathogen or set of related pathogens and/or immunogenic tick-saliva antigens (the trigger). This is followed by a systemic immune response that cross-reacts with host antigens on activated epithelium, for example via molecular mimicry. Stressed cholangiocytes upregulate certain surface molecules, becoming targets of periductal neutrophilic/Th17 inflammation. Gut–liver-axis mechanisms, microbial translocation, possible autoantigens and self-reinforcing inflammatory circuits sustain the disease (drivers) independently of the original exposure. Genetic susceptibility provides a Treg-deficient, Th17-favoring setting but is neither necessary nor sufficient.

One apparent objection to a tick-triggered model is that PSC responds biochemically to oral vancomycin, an antibiotic confined to the gut lumen. The characteristics of that response in fact support the trigger-driver separation rather than undermining it. The benefit is not PSC-specific: oral vancomycin induces or maintains remission in 55% of children with IBD and no PSC (Ancona 2025). It depends on luminal delivery, with vancomycin solution outperforming capsules for biochemical remission (adjusted odds ratio 23.1, 95% CI 2.11 to 253) (Raisanen 2025), which a systemically distributed target would not predict. It persists after colectomy, in the ileal pouch. And rifaximin, also non-absorbed but with a different spectrum, produces no change in alkaline phosphatase (+0.9%, p = 0.47). A luminal, gram-positive-directed antibiotic therefore acts on a luminal driver and is uninformative about the initiating trigger, which on our model is no longer present by the time disease is diagnosed.

This trigger–driver model describes a self-limited initiating infection that precipitates durable autoimmunity; it does not invoke persistent Borrelia infection or the contested entity of “chronic Lyme disease,” and the established disease is autoimmune rather than an active infection. This resolved-infection-to-autoimmunity paradigm is exemplified by the now-established causal link between Epstein–Barr virus and multiple sclerosis^49^, in which a common, self-limited infection triggers autoimmunity through molecular mimicry^50^.

That paradigm has since been applied to PSC itself, and the result bears directly on the present hypothesis. Deep T-cell receptor beta repertoire analysis identified 1,008 PSC-associated clonotypes significantly more abundant in PSC than in controls (p = 2.9 × 10-43) and further enriched in PSC liver relative to PSC blood; 332 of them map to Epstein-Barr virus, and EBV-specific monoclonal antibodies have been cloned from B cells in PSC livers (ElAbd 2025). This is the strongest available demonstration that an exogenous antigen drives a liver-enriched clonal response in PSC, and it is precisely the shape of mechanism our trigger-driver model proposes. It does not settle which antigen initiates the process. EBV seroprevalence is approximately 95% in adults and cannot by itself account for a greater than 30-fold geographic gradient in PSC, whereas Ixodes exposure varies by orders of magnitude across the same regions in which PSC incidence varies. The two are not mutually exclusive: EBV may act as a driver, a co-factor, or the dominant trigger in a subset of patients. The discriminating experiment is explicit and inexpensive. Published clonotype reference sets do not contain Anaplasma, Neoehrlichia, Babesia or Chlamydiales antigens; adding them would establish whether PSC-associated clonotypes also recognise Ixodes-borne agents, and a null result would materially weaken our hypothesis.

### Testable predictions

The hypothesis generates concrete, falsifiable predictions:

Epidemiologic. (1) PSC patients should report higher lifetime tick exposure than matched local controls. (2) As Ixodes ranges expand, PSC incidence should rise in newly endemic regions after a latency. (3) Within countries, PSC should cluster where Ixodes exposure is documented (e.g., along Australia’s I. holocyclus endemic eastern seaboard). A PSC cluster in a region without Ixodes-borne pathogen exposure or travel would substantially weaken the hypothesis. (4) Systematic magnetic resonance cholangiography screening of the IBD population in Alaska or Greenland should continue to find no PSC; finding screen-detected PSC at the rates reported elsewhere (8.1 to 9.0% of adults with IBD) would remove the strongest natural experiment supporting the hypothesis.

*Serologic*. PSC patients should show elevated antibodies to *Ixodes*-borne pathogen antigens versus matched controls (IBD-only, PBC, and healthy controls). Testing should include *Anaplasma*, *Neoehrlichia*, *Ehrlichia*, *Babesia*, *Rickettsia*, *Coxiella*, *Borrelia*, *Bartonella*, and genus-wide Chlamydiales and perhaps others, plus alpha-gal IgE. Direct detection of microbial nucleic acid in bile or liver would strongly support the hypothesis, though its absence would not refute a post-infectious mechanism.

Specimen choice should be specified in advance, because a negative result in the wrong compartment is uninformative. Enterococcus gallinarum was undetectable in stool yet recoverable from liver in the model in which it was causal (Manfredo Vieira 2018), and outer-membrane vesicles from Klebsiella pneumoniae and Proteus mirabilis reproduced hepatic injury in the absence of culturable organisms (Dorner 2024). Negative blood or stool studies therefore do not exclude a tissue-resident or vesicle-borne antigen; liver, bile and portal blood are the specimens of interest, and vesicle-directed as well as organism-directed assays should be used.

Assay design should control for Epstein-Barr virus and cytomegalovirus reactivity, because the two leading candidate triggers are confounded at the level of the assay rather than at the level of biology. Sera from patients with polymerase-chain-reaction-confirmed active EBV infection give false-positive anti-Borrelia results in 51.9% (IgM) and 63.0% (IgG) by indirect immunofluorescence and in 22.2% and 29.6% by enzyme immunoassay, most often directed against flagellin p41 and OspC, with lower but comparable rates for cytomegalovirus and BK virus (Wojciechowska-Koszko 2022). PSC patients carry an elevated Epstein-Barr antibody burden and a liver-enriched EBV-reactive T-cell repertoire (ElAbd 2025). A tick-pathogen serosurvey in PSC would therefore be expected to return positive results whether or not the vector-borne hypothesis is correct, and a positive result obtained without this control would be uninterpretable. Concurrent EBV and cytomegalovirus serology and viral-load measurement, reciprocal cross-adsorption of reactive sera, and confirmation of positives by an assay not based on whole-cell or flagellar antigen should be specified in the protocol in advance rather than invoked in response to a reviewer. The same constraint applies symmetrically: it is the reason we do not treat published tick-pathogen seroprevalence in hepatobiliary disease as evidence for the hypothesis.

Molecular (speculative). Anti-integrin αvβ6 autoantibodies are highly specific for PSC, but their prevalence is conditional on concomitant IBD. Sensitivity is 97.8% in PSC with ulcerative colitis versus 53.8% in PSC without IBD (Yasuda 2025), and in a four-cohort comparison anti-αvβ6 was raised in PSC-UC and PSC-Crohn’s relative to PSC alone (p < 0.0001 and p < 0.003) while PSC alone did not differ from healthy controls (Bloemen 2025). This conditionality sharpens the prediction rather than weakening it: integrin αvβ6 is selectively expressed on the disease-associated bile-duct and colonic epithelium, and if the autoantibody tracks colonic involvement then the colonic epithelium is the more likely primary site of cross-reactivity. Several tick-borne pathogens enter cells through integrin-binding proteins that share the αv subunit with integrin αvβ6. An immune response to such a pathogen could therefore cross-react with host αvβ6, on colonic epithelium first and on stressed cholangiocytes thereafter. Structural or serologic cross-reactivity between a pathogen integrin-binding domain and host αvβ6 should be explored, analysing PSC with and without IBD separately.

*Animal model (speculative)*. Chronic tick-pathogen exposures of mice with a vulnerable genetic background may reproduce a PSC-like cholangiopathy.

See further discussion in Supplementary Note S6.

### Limitations

The principal limitation of the evidence presented here is the ecological fallacy: region- or state-level correlations cannot demonstrate that individuals with PSC were exposed to ticks or tick-borne pathogens. The other major limitation is the paucity of PSC incidence and prevalence data worldwide.

Several other cautions apply:

Multiple comparisons. We examined 24 U.S. exposures. A Benjamini-Hochberg correction of the false-discovery rate across all 24 exposures leaves the three Ixodes-borne pathogens significant (q = 0.007, 0.007 and 0.007 for Anaplasma, Babesia and Powassan virus), whereas the Amblyomma-borne control (Ehrlichia chaffeensis) remains non-significant. Individual small-sample results, particularly the rarer Ixodes-borne exposures (Babesia, Powassan virus), still need replication. Powassan virus disease is the sparsest surface and non-reporting states were treated as zero incidence (Supplementary Note S1); we therefore also report the correlation with non-reporting states excluded rather than zero-filled [AUTHOR TO INSERT: partial r], since zero-filling a non-reporting state infers absence of disease from absence of surveillance.

Cross-country fragility. The PSC-Lyme rank correlation depends on region selection and Lyme case definition (Supplementary Note S4). The Netherlands illustrates this directly. We use a reported incidence of 40 per 100,000, whereas a recent systematic review places national Dutch Lyme incidence above 100 per 100,000 per year, alongside Belgium, Finland and Switzerland (Burn 2023). Substituting that value makes the Netherlands a high-exposure, low-PSC outlier. We therefore state the Lyme source for each region in Table 1 and report a sensitivity analysis substituting systematic-review values wherever they exist (Supplementary Note S4): [AUTHOR TO INSERT: rank correlation under substitution]. We regard this as the most fragile element of the cross-country comparison; the within-country analyses, which do not depend on Lyme surveillance at all, carry more weight.

#### MarketScan generalizability

The commercially insured population does not represent the U.S. census population; commercial-claims prevalence typically runs 2-4-fold below population-based registries, and is affected by interstate referral. The 2019 estimates are inflated by recoding of prevalent cases to the new code.

#### Ancestry adjustment

This adjustment is linear and ecological and cannot fully capture the HLA contribution; residual ancestry confounding cannot be excluded.

#### Lyme as proxy

Reported Lyme incidence derives from heterogeneous surveillance with varying case definitions.

#### Candidate pathogens

These have not been systematically tested in PSC cohorts; the pathogen discussion is speculative and requires prospective serologic and molecular validation. We cannot single out one agent. Indeed, an as yet unrecognized pathogen might be involved.

#### Exposure data quality

The tick-borne pathogen exposures were derived from CDC annual and cumulative surveillance data rather than the NNDSS weekly case tables, which undercount states that report annually (notably Minnesota); the derivation and its quality control are detailed in Supplementary Note S1.

#### Prevalence versus incidence

Our main U.S. pathogen results use PSC prevalence rather than incidence. Incidence would be a more meaningful measure for causation but is much noisier in these data.

#### Spatial clustering

PSC prevalence is geographically clustered. The *Anaplasma*-PSC association stays significant in spatial-lag and spatial-error regressions and after an effective-sample-size correction; however, the ancestry-only model still leaves significant clustering in its residuals, meaning some unmeasured factor that is itself geographically clustered could be contributing (Supplementary Note S4).

A comprehensive evaluation of all conceivable confounders is beyond any single ecological study.

## Conclusion

PSC has long been viewed as an idiopathic immune-mediated disease; the evidence assembled here, though ecological, supports reconsidering that view. PSC prevalence and incidence correlate with several markers of *Ixodes*-endemic environments in Europe and North America. This association is vulnerable to heterogeneous ascertainment, correlated regional exposures, residual spatial confounding, and the ecological fallacy, and it does not identify a causal vector or pathogen. The hypothesis is nonetheless falsifiable at the epidemiologic, serologic, molecular, and experimental levels. Confirmation would help identify at-risk populations and enable preventive or early-intervention strategies; refutation would narrow the search for an environmental trigger of PSC.

It is unlikely that every case of PSC is triggered by a tick bite. Ticks may represent one etiologic pathway, and other triggers may be pathobionts, metabolic products, or orally acquired organisms that share critical epitopes with the tick-borne agents. It seems likely that one or more of these factors might act as a “driver” of this disease after its initiation by the “trigger”. The biliary tree and colon present a complex antigenic surface, and several environmental agents and microbiome-related factors may converge to cause this complex disease.

## Abbreviations

ACS: American Community Survey
AIH: autoimmune hepatitis
AQS: Air Quality System (EPA)
ArboNET: National Arboviral Disease Surveillance System (CDC)
BRFSS: Behavioral Risk Factor Surveillance System (CDC)
CAPC: Companion Animal Parasite Council
CCAE: Commercial Claims and Encounters (MarketScan)
CD4 / CD8: T-cell markers
CDC: Centers for Disease Control and Prevention
CI: confidence interval
CLO: *Chlamydia*-like organism
DNA: deoxyribonucleic acid
DR3: HLA-DRB1*03 serotype
EPA: U.S. Environmental Protection Agency
GBIF: Global Biodiversity Information Facility
GWAS: genome-wide association study
HLA: human leukocyte antigen
IBD: inflammatory bowel disease
ICD-10-CM: International Classification of Diseases, 10th Revision, Clinical Modification
IDEXX: IDEXX Laboratories (veterinary diagnostics)
IgA/IgE: immunoglobulin A / E
IL-2: interleukin-2
IL-2Rα (CD25): interleukin-2 receptor α-chain
IRB: Institutional Review Board
LFTs: liver function tests
LPS: lipopolysaccharide
MDCR: Medicare Supplemental (MarketScan)
NF-κB: nuclear factor κB
NNDSS: National Notifiable Diseases Surveillance System (CDC)
OR: odds ratio
p-ANCA: perinuclear anti-neutrophil cytoplasmic antibody
PBC: primary biliary cholangitis
PM2.5: particulate matter ≤2.5 µm diameter
PNSP: Pesticide National Synthesis Project (USGS)
PSC: primary sclerosing cholangitis
Th17: T-helper-17 cell
Treg: regulatory T cell
USGS: U.S. Geological Survey
UV: ultraviolet
WONDER: Wide-ranging Online Data for Epidemiologic Research (CDC)

Gene symbols (IL2, IL2RA, REL, CARD9, MDR2, IL-10) follow standard nomenclature.

## Data Availability Statement

Analysis code will be archived at Zenodo with a citable DOI upon acceptance by a peer-reviewed journal, and will be made available by the corresponding author on request during peer review. Some of the key data is presented in the Supplement. The public exposure sources (CDC, USGS, EPA, U.S. Census Bureau) are cited in the Methods and Supplementary Methods.

## Author Contributions

Kevin M. Johnson: Conceptualization; Data curation; Formal analysis; Investigation; Methodology; Software; Validation; Visualization; Writing – original draft; Writing – review and editing. Declaration of generative AI and AI-assisted technologies in the writing process: During the preparation of this work, the author used Claude Opus 4.8 to organize the manuscript and its contents. The author reviewed and edited the output as needed and takes full responsibility for the content of the published article.

New references added in this revision (to be renumbered on acceptance)

## Supplement

### Supplementary Methods

#### S1. Data sources

This note describes the public data sources and the construction of the 24 state-level exposures as shown in Fig. 5 and Supplementary Table S4.

We measured *Ixodes* density as the occurrence-record density of *I. scapularis* and *I. pacificus* per 1,000 square miles, using records from the Global Biodiversity Information Facility (GBIF).

Survey effort biases this proxy, because it reflects where people recorded ticks rather than true questing density.

State-level incidence of the *Ixodes*-borne pathogens was taken from CDC annual and cumulative surveillance tables:

- *Anaplasma phagocytophilum* (anaplasmosis) and *Ehrlichia chaffeensis* (ehrlichiosis): CDC WONDER annual data (2018, 2019, 2021).
- Babesiosis: CDC annual babesiosis surveillance summary (2018–2019 counts).
- Powassan virus disease: CDC ArboNET annual data (2018–2021); non-reporting states treated as zero incidence.
- Lyme disease incidence: CDC WONDER annual data (2018–2022).

Counts were converted to average annual rates per 100,000 using 2020 Census state populations. We used three non-*Ixodes* tick-borne infections as within-class negative controls. *Amblyomma* transmits *Ehrlichia chaffeensis*, and *Dermacentor* and *Amblyomma* transmit spotted fever rickettsiosis and tularemia.^51^

Tick-borne disease incidence data are strongly right-skewed and zero-inflated: 27 of 36 states report fewer than 0.5 anaplasmosis cases per 100,000, separated by fractions of a case, while the upper range spans a hundred-fold gradient. We therefore analyze these as log10(x + 0.1), as we did for cross-country Lyme incidence (Figure 1).

For tick ecology we used deer density^52^ and forest cover.^53^ For climate and ultraviolet exposure we used state-centroid latitude. For agricultural and chemical rule-outs we used cumulative 1992 to 2017 application of aldicarb, total carbamates, organophosphates, and pyrethroids.^54^ For rural and water rule-outs we used reliance on private well water^55^ and percent rural population.^56^ For pollution rule-outs we used PM2.5 and ozone.^57^ For detection bias and reverse causation we used adult liver-transplant-center density^58^ and outpatient antibiotic prescribing.^59^ Two independent veterinary replicates provide canine *Anaplasma* seroprevalence^44^ and cat *Bartonella henselae* seroprevalence^45^. We also looked at adult current-smoker prevalence,^60^ because smoking is known to be inversely associated with PSC.

We used annual rather than weekly data. The NNDSS weekly tables report a year-to-date cumulative count at each week. A state that reports a given disease annually rather than weekly does not appear in the weekly stream. Minnesota is the most consequential case: it reports several tick-borne diseases annually and is endemic for all four *Ixodes*-borne pathogens studied here.

Supplementary Table S1. reports the effect of using annual rather than weekly data. All three *Ixodes*-borne correlations remained significant after correction, and the within-region (Northeast + Midwest) partials, adjusted for ancestry alone, are +0.81 to +0.85. The *Amblyomma*-borne control (*E. chaffeensis*) became more clearly dissociated. Correcting the data therefore sharpened the specificity pattern reported in the main text.

**Supplementary Table S1.** Effect of using annual rather than weekly NNDSS surveillance data on the *Ixodes*-borne exposure surfaces. Ancestry-adjusted partial correlations with state PSC prevalence. NE/MW denotes the Northeast and Midwest census-region block.

| Pathogen | Weekly-table value | Annual-data value | Nature of the weekly-table error |
|---|---|---|---|
| *A. phagocytophilum* | ancestry-partial +0.63 | +0.52 | Minnesota entered as 8 cases vs ~1,488 in WONDER annual (≈186-fold undercount) |
| Babesiosis | ancestry-partial +0.46 | +0.52 (+0.81 NE/MW) | endemic Minnesota dropped; New York over-counted ≈6-fold |
| Powassan virus | ancestry-partial +0.78 (n = 9) | +0.52 (n = 36; +0.85 NE/MW) | highest-burden state (Minnesota) undercounted (≈0.02/100k vs national maximum) |
| *E. chaffeensis* (*Amblyomma* control) | ancestry-partial −0.09 | −0.22 | same weekly-table source |

The *Dermacentor*/*Amblyomma*-borne controls, tularemia and spotted fever rickettsiosis, share the weekly-table source but are unaffected by the annual-reporter gap: their burden concentrates in central U.S. states that report weekly, and Minnesota is correctly absent (non-endemic) rather than dropped.<sup>51</sup> Their null-to-negative associations stand as reported.

## Supplementary Results

Two data tables open this section: a comparison of high versus zero PSC high-latitude populations (Table S2) and the state-level PSC, AIH, and PBC prevalence underlying the U.S. ecological analysis (Table S3).

**Supplementary Table S2.** High-latitude comparison of high-PSC versus zero-PSC populations. Among the factors examined, established human exposure to *Ixodes*-borne pathogens is the feature distinguishing high-PSC from zero-PSC populations.

| Factor | Finland / Scandinavia | Alaska Natives | Greenland |
|---|---|---|---|
| Northern latitude (>60°N) | Yes | Yes | Yes |
| Cold climate | Yes | Yes | Yes |
| Livestock / reindeer exposure | Yes | Yes | Yes |
| Rodent exposure | Yes | Yes | Yes |
| Waterborne illness burden | Yes | Yes | Yes |
| Autoimmune hepatitis present | Yes | Yes (very high) | Yes |
| Primary biliary cholangitis present | Yes | Yes | Yes |
| Inflammatory bowel disease present | Yes | Yes | Yes |
| Established *Ixodes*-borne pathogen exposure | Yes | No | No |
| Reported PSC | Yes | None | None |

**Supplementary Table S3.**
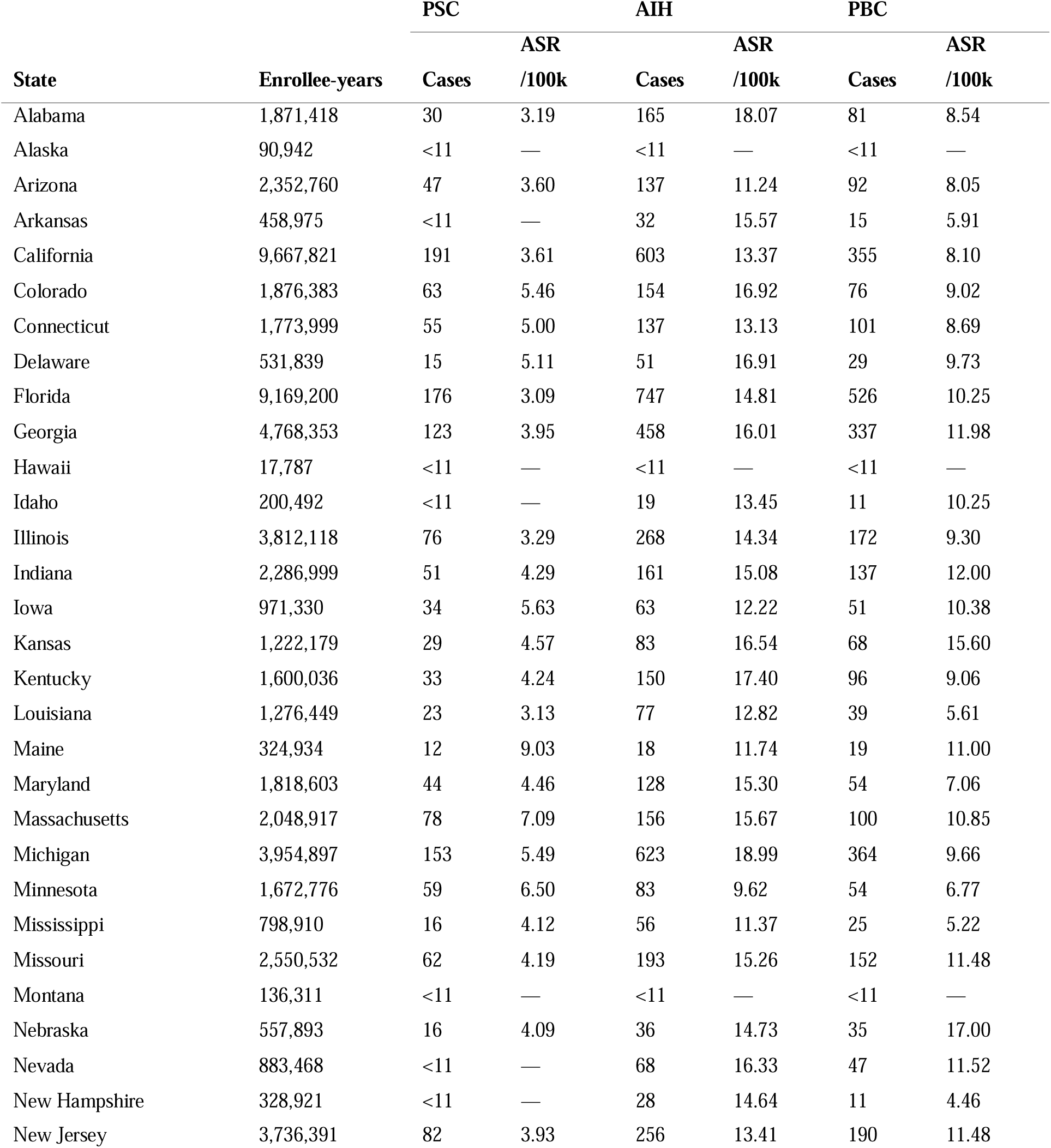
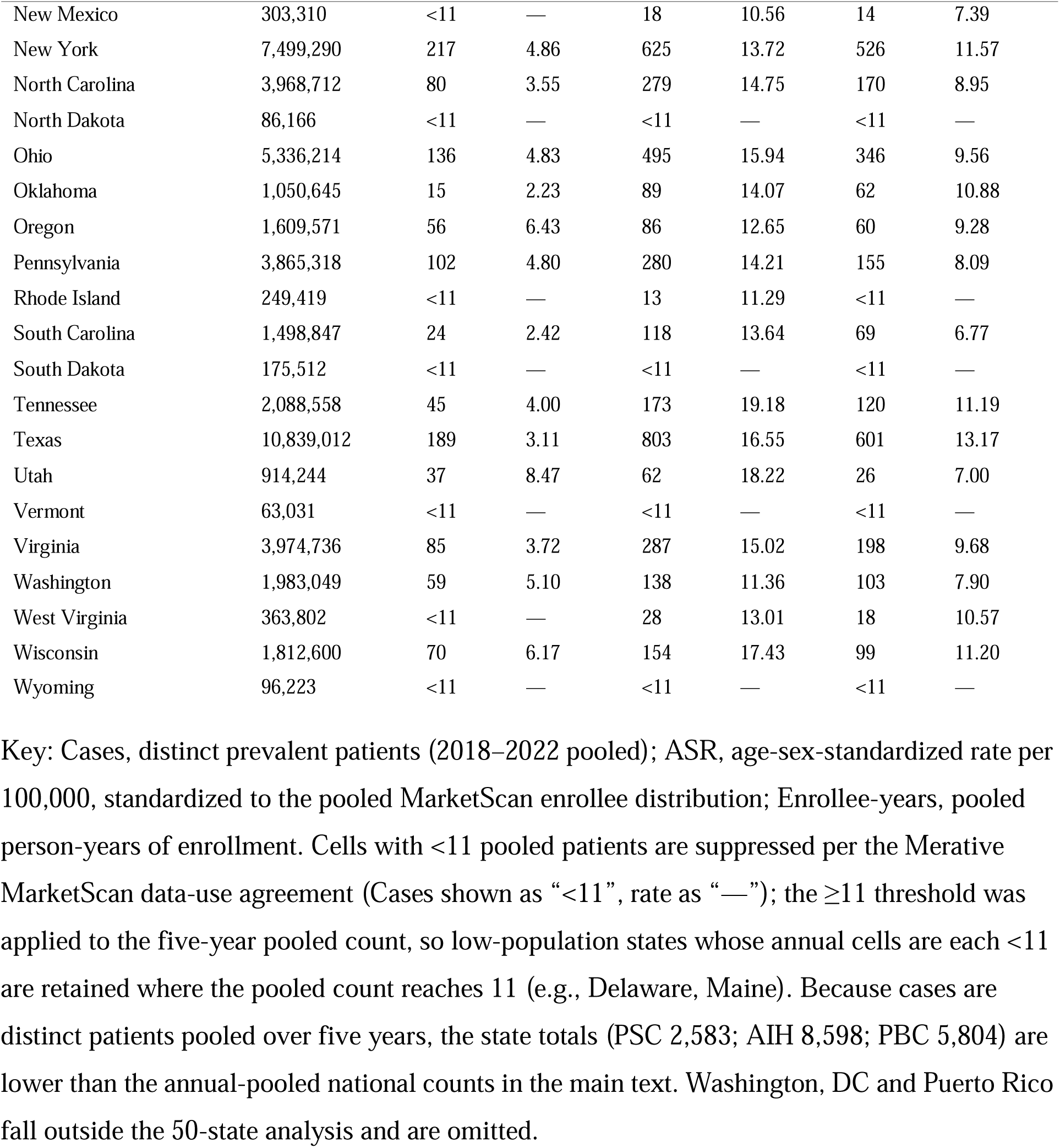
State-level prevalence of PSC, AIH, and PBC in MarketScan (50 U.S. states, 2018–2022 pooled). These are the state-level values underlying the U.S. ecological analysis (Figures 3–5). Distinct prevalent patients and age-sex-standardized prevalence per 100,000 for the 50 U.S. states, from Merative MarketScan (CCAE and MDCR), pooled 2018–2022.

### S2. Alternative environmental exposures

This note describes the findings for the 24 state-level exposures as shown in Fig. 5 and Supplementary Table S4.

#### Agricultural pesticides

In the U.S., after ancestry adjustment, no pesticide is positively associated with PSC at the state level. On the log scale used throughout, every pesticide runs negative (aldicarb r = −0.32; total carbamates −0.39; organophosphates −0.48; pyrethroids −0.44). *Ixodes* density and pathogens are positively associated with PSC while every pesticide is not, so a generic rural-agriculture confounder is unlikely.

The Mayo Clinic exposome studies found agricultural carbamates elevated in the bile of PSC patients, and the residential carbamate carbaryl reduced, with an odds ratio near 0.33.^61^ Although those biomarkers might mark rural residence and so stand in for outdoor exposures that increase tick contact, our data do not support that reading. Aldicarb geography follows Southeastern peanut, cotton, and citrus farming and is disjoint from *Ixodes* geography (r = −0.13), and PSC is, if anything, inversely associated with aldicarb. A dietary and hydrological explanation is simpler. Aldicarb and related residues reach consumers through the food supply and groundwater, largely independently of where farmers applied them, so a patient’s bile burden reflects diet and water source rather than residence.

#### Rural residence and water

Percent rural is inversely associated after ancestry adjustment (−0.33), and reliance on private well water attenuates to non-significance (+0.17, from a raw +0.37). Holding either variable fixed leaves the PSC-*Ixodes* partial correlation essentially unchanged.

#### Air pollution

PM2.5 (−0.31) and ozone (about 0.00) correlations run null to inverse.

#### Antibiotic prescribing

Outpatient antibiotic prescribing is strongly inversely associated (−0.59), which reflects the high-prescribing South against the low-prescribing Northeast and Pacific. Adjusting for it leaves the PSC-*Ixodes* partial unaffected. One reading consistent with the hypothesis is that heavy outpatient prescribing, which often includes doxycycline, blunts the sequelae of tick-borne infection. Ecological data cannot settle this.

#### Detection bias and reverse causation

Adult liver-transplant-center density is the most PSC-specific access variable and also the one most vulnerable to reverse causation. It is essentially uncorrelated with PSC (+0.07), and adjusting for it leaves the PSC-*Ixodes* partial unchanged (+0.380 against +0.381). Detection bias, in which more specialists produce more diagnoses, and reverse causation, in which more PSC attracts more specialists, both act through specialist density, so this single adjustment addresses both. Transplant density is also null against AIH (+0.13) and PBC (+0.01).

**Supplementary Table S4.** State-level ecological correlations of PSC prevalence with 24 exposure surfaces (MarketScan 2018–2022, ≥ 11-case states).

| Exposure | Category | n | r | r log† | ρ | 95% CI‡ | Source |
|---|---|---|---|---|---|---|---|
| *Anaplasma phagocytophilum* incidence | *Ixodes* pathogen | 36 | +0.52* | **+0.50**\* | +0.33 | +0.20, +0.72 | CDC WONDER |
| Babesiosis incidence | *Ixodes* pathogen | 31 | +0.52* | **+0.56**\* | +0.58* | +0.24, +0.76 | CDC WONDER |
| Powassan virus incidence | *Ixodes* pathogen | 36 | +0.52* | **+0.52**\* | +0.42* | +0.22, +0.73 | CDC ArboNET |
| *Ixodes* tick density | *Ixodes* habitat | 36 | +0.38* | **+0.26** | +0.38* | -0.08, +0.55 | GBIF; Eisen 2016 |
| Lyme disease incidence | *Ixodes* proxy | 36 | +0.40* | **+0.33** | +0.30 | -0.01, +0.59 | CDC WONDER |
| *Ehrlichia chaffeensis* (*Amblyomma*) | Non-*Ixodes* tick | 36 | -0.22 | **-0.26** | -0.21 | -0.55, +0.08 | CDC NNDSS |
| Spotted Fever Rickettsiosis | Non-*Ixodes* tick | 34 | -0.34 | **-0.40**\* | -0.37* | -0.66, -0.07 | CDC NNDSS |
| Tularemia | Non-*Ixodes* tick | 31 | -0.38* | **-0.39**\* | -0.02 | -0.66, -0.03 | CDC NNDSS |
| Canine *Anaplasma* seroprevalence | Vet replicate | 36 | **+0.49**\* | — | +0.52* | +0.19, +0.71 | CAPC |
| Cat *Bartonella* seroprevalence | Vet replicate | 36 | **-0.35**\* | — | -0.33* | -0.61, -0.02 | Osikowicz 2021 |
| Deer density | Tick ecology | 36 | **-0.19** | — | -0.06 | -0.49, +0.16 | Wildlife Informer |
| Forest cover % | Tick ecology | 36 | **+0.21** | — | +0.08 | -0.13, +0.51 | USDA FIA 2016 |
| Latitude | Climate/UV | 36 | **+0.55**\* | — | +0.72* | +0.27, +0.75 | State centroid |
| Adult smoking prevalence | Data-source check | 36 | **-0.69**\* | — | -0.62* | -0.83, -0.46 | CDC BRFSS |
| Aldicarb (cumulative) | Pesticide | 33 | -0.19 | **-0.32** | -0.39* | -0.60, +0.03 | Wieben 2019 |
| Total agricultural carbamates | Pesticide | 36 | -0.08 | **-0.39**\* | -0.19 | -0.64, -0.06 | Wieben 2019 |
| Organophosphates | Pesticide | 36 | -0.27 | **-0.48**\* | -0.19 | -0.70, -0.17 | Wieben 2019 |
| Pyrethroids | Pesticide | 36 | -0.18 | **-0.44**\* | -0.18 | -0.68, -0.13 | Wieben 2019 |
| Private well water % | Water | 36 | **+0.17** | — | +0.16 | -0.17, +0.48 | Dieter 2015 |
| Rural population % | Rural | 36 | **-0.33** | — | -0.53* | -0.59, +0.01 | U.S. Census |
| PM2.5 | Air pollution | 36 | **-0.31** | — | -0.32 | -0.59, +0.02 | EPA AQS |
| Ozone | Air pollution | 36 | **-0.00** | — | -0.04 | -0.34, +0.33 | EPA AQS |
| Liver transplant centers / M | Care access | 36 | **+0.07** | — | +0.13 | -0.27, +0.39 | Am. Liver Foundation |
| Outpatient antibiotics / 1000 | Care access | 36 | **-0.59**\* | — | -0.65* | -0.77, -0.32 | CDC (IQVIA) |

All correlations except the two veterinary replicates are adjusted for European-ancestry fraction; asterisks mark p < 0.05. Columns: r is the untransformed Pearson partial; r log is the Pearson partial on the log10(x + 0.1) scale; ρ is the Spearman partial. † 95% CI applies to the primary correlation estimates (bolded).

**Supplementary Table S5.** Italy regional Lyme borreliosis case-rate proxy. ^14^ Regional Lyme values used in the within-Italy analysis (Figure 6,). This is a center-ascertained case-rate proxy, not population-based surveillance incidence. Cases were drawn from Trevisan et al. 2023, a multi-center clinical series (eight hospital centers, five regions, January 2010– August 2022, 12.67 years); annualized rate = cumulative rate ÷ 12.67.

| Region | Lyme cases | Population | Cumulative /100,000 | Annualized /100,000/yr |
|---|---|---|---|---|
| Friuli-Venezia Giulia | 517 | 1,194,095 | 43.30 | 3.420 |
| Liguria | 62 | 1,509,908 | 4.11 | 0.324 |
| Veneto | 120 | 4,851,851 | 2.47 | 0.195 |
| Valle d’Aosta | 3 | 122,714 | 2.44 | 0.193 |
| Emilia-Romagna | 95 | 4,465,678 | 2.13 | 0.168 |
| Lombardia | 197 | 10,035,481 | 1.96 | 0.155 |
| Piemonte | 65 | 4,255,702 | 1.53 | 0.121 |
| Toscana | 51 | 3,660,834 | 1.39 | 0.110 |
| Trentino-Alto Adige* | 15 | 1,086,095 | 1.38 | 0.109 |
| Molise† | 3 | 287,966 | 1.04 | 0.082 |
| Marche | 9 | 1,481,252 | 0.61 | 0.048 |
| Sardegna | 8 | 1,561,339 | 0.51 | 0.040 |
| Campania | 27 | 5,575,025 | 0.48 | 0.038 |
| Lazio | 27 | 5,710,272 | 0.47 | 0.037 |
| Umbria | 4 | 851,954 | 0.47 | 0.037 |
| Puglia | 16 | 3,874,166 | 0.41 | 0.033 |
| Abruzzo | 4 | 1,268,430 | 0.32 | 0.025 |
| Calabria | 5 | 1,832,147 | 0.27 | 0.022 |
| Sicilia | 12 | 4,779,371 | 0.25 | 0.020 |
| Basilicata | 0 | 529,897 | 0.00 | 0.000 |

\* Trevisan reports Trentino-Alto Adige combined; in the analysis this rate is applied to both the Bolzano and Trento autonomous provinces (n = 20 analysis units). † Molise is excluded from the correlation (no PSC incidence estimate).

**Supplementary Table S6.** Japanese prefecture-level PSC prevalence and tick density. PSC point prevalence (per 100,000; national designated-intractable-disease certifications, personal communication A. Tanaka January 2026). Ixodes spp. and Haemaphysalis longicornis relative density (0–3 ordinal, from ECDC-style distribution maps and published surveys) for all 47 prefectures. Combined = Ixodes + H. longicornis (0–6).

| Prefecture | PSC prevalence (/100,000) | Ixodes density (0–3) | H. longicornis density (0–3) | Combined (0–6) |
|---|---|---|---|---|
| Tottori | 2.07 | 2 | 2 | 4 |
| Nara | 1.95 | 1 | 2 | 3 |
| Akita | 1.78 | 3 | 2 | 5 |
| Iwate | 1.75 | 3 | 2 | 5 |
| Ehime | 1.57 | 1 | 3 | 4 |
| Ishikawa | 1.55 | 2 | 2 | 4 |
| Saga | 1.53 | 1 | 2 | 3 |
| Fukui | 1.49 | 2 | 2 | 4 |
| Tokushima | 1.45 | 2 | 2 | 4 |
| Gunma | 1.43 | 2 | 2 | 4 |
| Gifu | 1.36 | 2 | 2 | 4 |
| Toyama | 1.30 | 2 | 2 | 4 |
| Hyogo | 1.28 | 1 | 2 | 3 |
| Nagasaki | 1.20 | 1 | 2 | 3 |
| Osaka | 1.16 | 1 | 2 | 3 |
| Kyoto | 1.15 | 2 | 2 | 4 |
| Wakayama | 1.14 | 1 | 2 | 3 |
| Fukuoka | 1.13 | 1 | 3 | 4 |
| Kanagawa | 1.10 | 1 | 2 | 3 |
| Saitama | 1.09 | 1 | 2 | 3 |
| Hokkaido | 1.09 | 3 | 1 | 4 |
| Chiba | 1.05 | 1 | 2 | 3 |
| Hiroshima | 1.03 | 1 | 2 | 3 |
| Miyagi | 1.02 | 2 | 2 | 4 |
| Tokyo | 0.99 | 1 | 1 | 2 |
| Aichi | 0.95 | 1 | 2 | 3 |
| Mie | 0.93 | 1 | 2 | 3 |
| Okayama | 0.92 | 2 | 2 | 4 |
| Fukushima | 0.92 | 2 | 2 | 4 |
| Ibaraki | 0.89 | 1 | 2 | 3 |
| Nagano | 0.85 | 2 | 2 | 4 |
| Kagoshima | 0.85 | 1 | 3 | 4 |
| Okinawa | 0.82 | 0 | 0 | 0 |
| Tochigi | 0.80 | 2 | 2 | 4 |
| Shimane | 0.79 | 2 | 2 | 4 |
| Yamaguchi | 0.78 | 2 | 3 | 5 |
| Shiga | 0.78 | 2 | 2 | 4 |
| Kochi | 0.77 | 2 | 2 | 4 |
| Yamanashi | 0.76 | 2 | 2 | 4 |
| Shizuoka | 0.71 | 2 | 3 | 5 |
| Kumamoto | 0.65 | 1 | 3 | 4 |
| Oita | 0.65 | 1 | 2 | 3 |
| Aomori | 0.52 | 3 | 2 | 5 |
| Yamagata | 0.50 | 2 | 2 | 4 |
| Miyazaki | 0.39 | 1 | 3 | 4 |
| Kagawa | 0.33 | 2 | 2 | 4 |
| Niigata | 0.29 | 2 | 2 | 4 |

Spearman correlation with PSC prevalence: Ixodes ρ = −0.02; H. longicornis ρ = −0.15; combined ρ = −0.13 (all p > 0.3, n = 47).

### S3. Temporal concordance

Where time series exist, tick range and PSC incidence have risen together. In the United States the *I. scapularis* range expanded from the 1970s, and PSC incidence in Minnesota rose over the same period.^7,62–63^ Scandinavian reports describe parallel increases in *Ixodes ricinus* abundance and in PSC incidence.^64–65^ Temporal concordance is weak evidence on its own, because many exposures have risen over the same decades, but it is consistent with the hypothesis.

### S4. Robustness and sensitivity analyses

#### Cross-country correlation

The cross-country PSC-Lyme rank correlation ranges from ρ = 0.71 (all 13 regions) to 0.87 (the 11 *Ixodes*-endemic regions), depending on region selection and Lyme case-definition. The PSC incidence for Sweden was adjusted as follows: the Bergquist et al. study included rates from 2002–2005 overlapping with the earlier Lindkvist et al. study. This revealed that the more inclusive ICD-10-based methodology in Bergquist et al. counted approximately 1.8 times more cases than the stricter diagnostic criteria used by Lindkvist et al during those same overlapping years. To enable direct comparison across studies, the Bergquist PSC incidence was adjusted from 3.3 to 1.8 per 100,000 to align with the Lindkvist methodology. Bergquist 2026 identifies PSC through the Swedish National Patient Register using broad ICD-10 codes (K83.0A, or cholangitis K83.0 + IBD) and reports crude incidence rising from 2.2 (2002) to 3.3 (2020) per 100,000. Lindkvist 2010 instead validated all 199 cases against Mayo criteria by chart review and got a crude incidence of 1.22 per 100,000. In any case, using the unadjusted Sweden figure instead still preserves the cross-region rank correlation, Spearman ρ = 0.87.

#### Spatial clustering

High-PSC states tend to sit next to other high-PSC states; the disease is geographically clustered rather than scattered at random. We measured this with Moran’s I, an index running from 0 (no spatial pattern) toward 1 (strong clustering); for PSC it was about 0.39–0.40 (p ≤ 0.002). Because neighboring states are not statistically independent, clustering like this can by itself inflate an apparent correlation, so we asked whether the Anaplasma-PSC association was merely a by-product of it. It was not. Clustering effectively shrinks the number of independent observations (our 36 states behave like roughly 29 truly independent ones) and after correcting for this the ancestry-adjusted association remained significant (effective n ≈ 29, p ≈ 0.002). How much clustering distorts a correlation depends on how strongly *both* maps are clustered at once; anaplasmosis is only moderately clustered (I ≈ 0.27), and two moderate patterns distort far less than one strong one. We also refit the association using two standard models that explicitly account for the influence of neighboring states, a spatial-lag and a spatial-error model, and, adjusting for European-ancestry fraction, the Anaplasma effect stayed significant in both (β = +0.06; p = 0.004 and 0.007). One caveat remains: after adjusting for ancestry alone, PSC was still significantly clustered (Moran’s I ≈ 0.40, p = 0.001), so we cannot exclude an unmeasured risk factor that is itself geographically clustered.

#### Do PSC and exposure hotspots overlap geographically?

Beyond whether each map is clustered on its own, we asked whether high-PSC states sit next to high-*exposure* states. A neighbor-based concordance measure bivariate Moran’s I (does a high-PSC state border high-exposure states?) was positive for all three Ixodes-borne exposures: anaplasmosis +0.25, Lyme +0.12, and Ixodes tick habitat +0.53. Smoothing each map by averaging every state with its neighbors, PSC tracked the three exposures at +0.63, +0.28, and +0.57. Both measures depend on how “neighbor” is defined, and both hinge on one influential state: Maine is the single strongest shared PSC/exposure hotspot, but it borders only New Hampshire, which is suppressed and out of sample. Under a strict definition that counts only shared land borders, Maine has no in-sample neighbor and drops out of the neighbor-based calculation entirely, which then falls to about +0.10–0.12 for all three exposures. That fragility is specific to the neighbor-based measure. When we instead simply remove Maine and recompute the ordinary state-to-state correlations, the associations for anaplasmosis and tick habitat are essentially unchanged (r = +0.48 and +0.34), whereas Lyme weakens sharply (+0.46 → +0.17) so only the Lyme association leans heavily on Maine.

#### Reproducibility

These analyses are reproduced by the MATLAB analysis code, which will be released as described in the Data Availability Statement.

## Supplementary Discussion

### S5. Additional candidate pathogens

This note expands the main text’s candidate-pathogen discussion (Candidate triggering pathogens; Table 2), reviewing the biliary/hepatic evidence and geographic fit for each candidate agent. It should be kept in mind that an as yet unrecognized agent could be involved.

#### Anaplasmataceae

Human granulocytic anaplasmosis causes substantial hepatic involvement, with periportal lymphohistiocytic infiltrates and elevated liver enzymes in roughly two-thirds of cases,^66^ and the injury appears immune-mediated rather than directly bacterial.^67^ Its geography also fits. In Europe, *A. phagocytophilum* prevalence in questing ticks and reservoir hosts is high (∼20%; 19.9% in Europe^68^). In North America, anaplasmosis was first recognized in Minnesota and Wisconsin (1994), a PSC epicenter where reported human incidence remains highest.^69^ Large Scandinavian surveys rank *A. phagocytophilum* and Candidatus *Neoehrlichia mikurensis* among the most prevalent *I. ricinus* pathogens.^70^ In southern Norway *N. mikurensis* reaches up to 25% of questing *I. ricinus* ticks.^71^ *N. mikurensis* is mechanistically suggestive. It has a tropism for vascular endothelium. Investigators have cultured it in human endothelial cells and seen it inside circulating endothelial cells,^79^ and it causes thrombotic and vascular disease.^80^ The biliary tree depends on the peribiliary vascular plexus for its blood supply, so endothelial injury in that plexus offers a plausible route to bile-duct damage. Serology for *N. mikurensis* in PSC cohorts would test this directly.

#### *Chlamydia*-like organisms (CLOs)

Ponsioen et al. reported elevated anti-*Chlamydia*-LPS IgA in PSC (≈80% of patients positive; OR 6.7, 95% CI 3.0–17.0), among the strongest serological signals linking any pathogen class to PSC.^72^ There was no species-specific *Chlamydia trachomatis* or *Chlamydia pneumoniae* signal, suggesting reactivity to a broader Chlamydiales epitope. Finnish ticks carry Chlamydiales at 40%, the highest reported anywhere, with evidence of tick-to-human transmission: 68% of tick-bite skin biopsies contained Chlamydiales DNA, 95– 98% similar to tick-derived organisms.^73^ This direct-transmission evidence comes from a country with one of the world’s highest PSC incidences. The Ponsioen finding has not been replicated with contemporary methods and should be interpreted cautiously given assay cross-reactivity. Although Chlamydiales are prevalent and diverse in European *I. ricinus*,^73–74^ they have never been systematically screened in *I. scapularis* or *Haemaphysalis longicornis*, a high-priority evidence gap.

#### *Bartonella*, which the ecological data argue against

*Bartonella* species show biliary tropism. Cats infected with *B. henselae* develop a lymphocytic cholangitis that resembles PSC, often with concurrent IBD,^75^ and childhood cat ownership has been reported as a risk factor for pediatric autoimmune liver disease.^76^ Our cat-*Bartonella* analysis (Figure 5; veterinary replicate) nevertheless found an inverse state-level association with PSC. *Bartonella* is also largely absent from Swedish *Ixodes ricinus* ticks^77^ and from Norwegian cats.^78^ We therefore think *Bartonella* is unlikely to be the primary trigger at the population scale, although it may contribute in specific settings.

#### *Borrelia*, which we exclude as the sole trigger

The cross-country correlation between PSC and Lyme is strong, but *Borrelia* cannot be the sole trigger. PSC occurs where classical Lyme borreliosis is absent or rare, including Japan, Australia, and New Zealand, and within the United States PSC tracks reported Lyme incidence only weakly and non-specifically, in contrast to *Anaplasma*. Lyme incidence is best read here as a proxy for living in an *Ixodes*-endemic environment rather than as the agent itself. Post-infectious phenomena such as antibiotic-refractory Lyme arthritis nonetheless show that an *Ixodes*-borne infection can start durable immune-mediated disease.

### S6. Mechanism: molecular mimicry, integrins, and the Treg and Th17 axis

This note expands the mechanisms suggested in the main text (Discussion).

#### Integrin cross-reactivity (hypothesized)

Anti-integrin αvβ6 autoantibodies occur in 73 to 91% of PSC patients across Japanese and Western cohorts and correlate with disease severity.^81–84^ Integrin αvβ6 is selectively expressed on the bile-duct and colonic epithelium that the disease targets. Several tick-borne pathogens use integrin-binding proteins to enter cells. *B. burgdorferi* P66 targets αvβ3, and *Bartonella* targets β1. The αv subunit is shared between these pathogen targets and the PSC autoantigen, and the β3 and β6 subunits share about 56% homology. An immune response to a pathogen integrin-binding domain could therefore cross-react with host αvβ6 on stressed cholangiocytes. Structural alignment and cross-adsorption would test this prediction. Shared αv usage and partial β-subunit homology are necessary but not sufficient for mimicry and do not by themselves establish cross-reactivity.

#### Experimental models

The largest common genetic contribution to PSC is the HLA-B*08:01 and DRB1*03:01 haplotype, which favors pro-inflammatory Th17 responses. The PSC and ulcerative-colitis risk loci define the IL-2, Treg and NF-κB axis.^85^ These include IL2, IL2RA, REL, and CARD9, Indeed, the IL-2Rα-knockout mouse spontaneously develops portal inflammation with biliary-ductular damage that resembles early human autoimmune cholangitis, together with an IBD-like colitis. Treg deficiency drives both. The cholangitis depends on CD8 cells, while the colitis depends on CD4 cells.^86^ A similar division has been observed in human PSC.

*Anaplasma phagocytophilum* produces a self-limited periportal hepatitis in immunocompetent mice, but in persistent-infection models it progresses to granulomatous hepatitis with a ductular reaction, a cholestatic biochemical pattern and periductal fibrosis. Because the organism does not infect cholangiocytes, the immune response mediates this bile-duct injury through Th17 and neutrophilic pathways. That matches human PSC immunology, including neutrophil extracellular traps and Th17 responses, and it matches the Th17-favoring genetic background.

These two observations motivate an animal experiment: low-dose *A. phagocytophilum, Ehrlichia muris* or other tick-borne pathogen or tick whole extracts exposure of IL-2Rα-knockout mice (possibly a heterozygous version since the homozygous mice are fragile), with cholangiography, alkaline phosphatase, and bile-duct histology as endpoints.

## References

1. Karlsen TH, Folseraas T, Thorburn D, Vesterhus M. Primary sclerosing cholangitis – a comprehensive review. J Hepatol. 2017;67(6):1298–1323. 10.1016/j.jhep.2017.07.022

2. Jiang X, Karlsen TH. Genetics of primary sclerosing cholangitis and pathophysiological implications. Nat Rev Gastroenterol Hepatol. 2017;14(5):279–295. 10.1038/nrgastro.2016.154

3. Andersen IM, Tengesdal G, Lie BA, et al. Effects of coffee consumption, smoking, and hormones on risk for primary sclerosing cholangitis. Clin Gastroenterol Hepatol. 2014;12(6):1019–1028. 10.1016/j.cgh.2013.09.024

4. Trivedi PJ, Bowlus CL, Yimam KK, et al. Epidemiology, natural history, and outcomes of primary sclerosing cholangitis: a systematic review of population-based studies. Clin Gastroenterol Hepatol. 2022;20(8):1687–1700. 10.1016/j.cgh.2021.08.039

5. Cooper J, Markovinovic A, Coward S, et al. Incidence and prevalence of primary sclerosing cholangitis: a meta-analysis of population-based studies. Inflamm Bowel Dis. 2024;30(11):2019–2026. 10.1093/ibd/izad276

6. Barner-Rasmussen N, Pukkala E, Jussila A, et al. Epidemiology, risk of malignancy and patient survival in primary sclerosing cholangitis: a population-based study in Finland. Scand J Gastroenterol. 2020;55(1):74–81. 10.1080/00365521.2019.1707277

7. Bakhshi Z, Hilscher MB, Gores GJ, et al. An update on primary sclerosing cholangitis epidemiology, outcomes and quantification of alkaline phosphatase variability in a population-based cohort. J Gastroenterol. 2020;55(5):523–532. 10.1007/s00535-020-01663-1

8. Toy E, Balasubramanian S, Selmi C, Li CS, Bowlus CL. The prevalence, incidence and natural history of primary sclerosing cholangitis in an ethnically diverse population. BMC Gastroenterol. 2011;11:83. 10.1186/1471-230X-11-83

9. Hurlburt KJ, McMahon BJ, Deubner H, et al. Prevalence of autoimmune liver disease in Alaska Natives. Am J Gastroenterol. 2002;97(9):2402–2407. 10.1016/S0002-9270(02)04376-9

10. Gantzel RH, Bagge CN, Villadsen GE, et al. The prevalence and disease course of autoimmune liver diseases in Greenland. Int J Circumpolar Health. 2024;83:2327693. 10.1080/22423982.2024.2327693

11. Dyson JK, Blain A, Foster Shirley MD, et al. Geo-epidemiology and environmental co-variate mapping of primary biliary cholangitis and primary sclerosing cholangitis. JHEP Rep. 2021;3(1):100202. 10.1016/j.jhepr.2020.100202

12. Gonzalez-Galarza FF, McCabe A, Santos EJMD, et al. Allele Frequency Net Database (AFND) 2020 update: gold-standard data classification, open access genotype data and new query tools. Nucleic Acids Res. 2020;48(D1):D783–D788. 10.1093/nar/gkz1029

13. Carbone M, Kodra Y, Rocchetti A, et al. Primary sclerosing cholangitis: burden of disease and mortality using data from the national rare diseases registry in Italy. Int J Environ Res Public Health. 2020;17:3095. 10.3390/ijerph17093095

14. Trevisan G, Ruscio M, Cinco M, et al. The history of Lyme disease in Italy and its spread in the Italian territory. Front Pharmacol. 2023;14:1128142. 10.3389/fphar.2023.1128142

15. Goto K, Kato H, Kanesaki M, et al. Epidemiology of Lyme disease, a growing tick-borne disease of concern, in Japan from May 2013 to March 2024: a descriptive study. IJID Reg. 2026;19:100919. 10.1016/j.ijregi.2026.100919

16. Puustinen L, et al. Incidence, prevalence, and causes of death of patients with autoimmune hepatitis: a nationwide register-based cohort study in Finland. Dig Liver Dis. 2019;51(9):1294–1299. 10.1016/j.dld.2019.01.015

17. Bergquist A, Kachru N, Aldvén M, et al. Burden of primary sclerosing cholangitis in Sweden (2002–2020): incidence, outcomes, healthcare utilization, and costs. Hepatol Commun. 2025;10(1):e0858. 10.1097/hc9.0000000000000858

18. Werner M, Prytz H, Ohlsson B, et al. Epidemiology and the initial presentation of autoimmune hepatitis in Sweden: a nationwide study. Scand J Gastroenterol. 2008;43(10):1232–1240. 10.1080/00365520802130183

19. Boberg KM, Aadland E, Jahnsen J, Raknerud N, Stiris M, Bell H. Incidence and prevalence of primary biliary cirrhosis, primary sclerosing cholangitis, and autoimmune hepatitis in a Norwegian population. Scand J Gastroenterol. 1998;33(1):99–103. 10.1080/00365529850166284

20. Nielsen KR, Midjord J, Johannesen HL, Grønbæk H. A nationwide study of autoimmune liver diseases in the Faroe Islands: incidence, prevalence, and causes of death 2004–2021. Int J Circumpolar Health. 2023;82(1):2221368. 10.1080/22423982.2023.2221368

21. Gudnason HO, Kristinsson JO, Bergmann OM, et al. Primary sclerosing cholangitis in Iceland 1992–2012. Laeknabladid. 2019;105(9):371–376.

22. Valgeirsson KB, Hreinsson JP, Björnsson ES. Increased incidence of autoimmune hepatitis is associated with wider use of biological drugs. Liver Int. 2019;39(12):2341–2349. 10.1111/liv.14224

23. Baldursdottir TR, Bergmann OM, Jonasson JG, Ludviksson BR, Axelsson TA, Björnsson ES. The epidemiology and natural history of primary biliary cirrhosis: a nationwide population-based study. Eur J Gastroenterol Hepatol. 2012;24(7):824–830. 10.1097/meg.0b013e328353753d

24. Liang H, Manne S, Shick J, Lissoos T, Dolin P. Incidence, prevalence, and natural history of primary sclerosing cholangitis in the United Kingdom. Medicine (Baltimore). 2017;96(24):e7116. 10.1097/md.0000000000007116

25. Webb GJ, Ryan RP, Marshall TP, Hirschfield GM. The epidemiology of UK autoimmune liver disease varies with geographic latitude. Clin Gastroenterol Hepatol. 2021;19(12):2587–2596. 10.1016/j.cgh.2021.01.029

26. Metcalf JV, Bhopal RS, Gray J, Howel D, James OF. Incidence and prevalence of primary biliary cirrhosis in the city of Newcastle upon Tyne, England. Int J Epidemiol. 1997;26(4):830–836. 10.1093/ije/26.4.830

27. Boonstra K, Weersma RK, van Erpecum KJ, et al. Population-based epidemiology, malignancy risk, and outcome of primary sclerosing cholangitis. Hepatology. 2013;58(6):2045–2055. 10.1002/hep.26565

28. van Gerven NM, Verwer BJ, Witte BI, et al. Epidemiology and clinical characteristics of autoimmune hepatitis in the Netherlands. Scand J Gastroenterol. 2014;49(10):1245–1254. 10.3109/00365521.2014.946083

29. Marzioni M, Bassanelli C, Ripellino C, Urbinati D, Alvaro D. Epidemiology of primary biliary cholangitis in Italy: evidence from a real-world database. Dig Liver Dis. 2019;51(5):724–729. 10.1016/j.dld.2018.11.008

30. Leung KK, Li W, Hansen B, et al. Primary sclerosing cholangitis–inflammatory bowel disease: epidemiology, mortality, and impact of diagnostic sequence. JHEP Rep. 2025;7(3):101272 (epub 2024). 10.1016/j.jhepr.2024.101272

31. Kim WR, Lindor KD, Locke GR 3rd, et al. Epidemiology and natural history of primary biliary cirrhosis in a US community. Gastroenterology. 2000;119(6):1631–1636. 10.1053/gast.2000.20197

32. Benson-Pope S, Stedman C, Ngu J. Epidemiology of primary sclerosing cholangitis in Canterbury, New Zealand: a population-based study. J Hepatol. 2025;82(S1):S70–S858 (EASL Congress 2025 abstract THU-358-YI). 10.1016/s0168-8278(25)01029-3

33. Ngu JH, Bechly K, Chapman BA, et al. Population-based epidemiology study of autoimmune hepatitis: a disease of older women? J Gastroenterol Hepatol. 2010;25(10):1681–1686. 10.1111/j.1440-1746.2010.06384.x

34. Ngu JH, Gearry RB, Wright AJ, Stedman CA. Low incidence and prevalence of primary biliary cirrhosis in Canterbury, New Zealand: a population-based study. Hepatol Int. 2012;6(4):796–800. 10.1007/s12072-011-9329-0

35. Lv T, Chen S, Li M, et al. Regional variation and temporal trend of primary biliary cholangitis epidemiology: a systematic review and meta-analysis. J Gastroenterol Hepatol. 2021;36(6):1423–1434. 10.1111/jgh.15329

36. Gazda J, Drazilova S, Janicko M, et al. The epidemiology of primary biliary cholangitis in European countries: a systematic review and meta-analysis. Can J Gastroenterol Hepatol. 2021;2021:9151525. 10.1155/2021/9151525

37. Mosites E, Miernyk K, Priest JW, et al. Giardia and Cryptosporidium antibody prevalence and correlates of exposure among Alaska residents, 2007–2008. Epidemiol Infect. 2018;146(7):888–894. 10.1017/s095026881800078x

38. Laaksonen M, Klemola T, Feuth E, et al. Tick-borne pathogens in Finland: comparison of *Ixodes ricinus* and *I. persulcatus* in sympatric and parapatric areas. Parasit Vectors. 2018;11:556. 10.1186/s13071-018-3131-y

39. Uusitalo R, Siljander M, Lindén A, et al. Predicting habitat suitability for *Ixodes ricinus* and *Ixodes persulcatus* ticks in Finland. Parasit Vectors. 2022;15:310. 10.1186/s13071-022-05410-8

40. Sievänen T, Neuvonen M, Pouta E, et al. Outdoor recreation and nature tourism in Finland: a national assessment. Metlan työraportteja. 2011;212:1–74.

41. van Schoor NM, Lips P. Worldwide vitamin D status. Best Pract Res Clin Endocrinol Metab. 2011;25(4):671–680. 10.1016/j.beem.2011.06.007

42. Jääskeläinen T, Itkonen ST, Lundqvist A, et al. The positive impact of general vitamin D food fortification policy on vitamin D status in a representative adult Finnish population: evidence from an 11-y follow-up based on standardized 25-hydroxyvitamin D data. Am J Clin Nutr. 2017;105(6):1512–1520. 10.3945/ajcn.116.151415

43. Lips P, Cashman KD, Lamberg-Allardt C, et al. Current vitamin D status in European and Middle East countries and strategies to prevent vitamin D deficiency: a position statement of the European Calcified Tissue Society. Eur J Endocrinol. 2019;180(4):P23–P54. 10.1530/EJE-18-0736

44. Companion Animal Parasite Council (CAPC). Anaplasma spp. canine seroprevalence maps, 2022 (IDEXX/Antech diagnostic data).

45. Osikowicz LM, Horiuchi K, Goodrich I, et al. Exposure of domestic cats to three zoonotic *Bartonella* species in the United States. Pathogens. 2021;10(3):354. 10.3390/pathogens10030354

46. Gofton AW, Doggett S, Ratchford A, et al. Bacterial profiling reveals novel ‘Ca. *Neoehrlichia*’, *Ehrlichia*, and *Anaplasma* species in Australian human-biting ticks. PLoS One. 2015;10(12):e0145449. 10.1371/journal.pone.0145449

47. Carpi G, Cagnacci F, Wittekindt NE, et al. Metagenomic profile of the bacterial communities associated with *Ixodes ricinus* ticks. PLoS One. 2011;6(10):e25604. 10.1371/journal.pone.0025604

48. Moutailler S, Valiente Moro C, Vaumourin E, et al. Co-infection of ticks: the rule rather than the exception. PLoS Negl Trop Dis. 2016;10(3):e0004539. 10.1371/journal.pntd.0004539

49. Bjornevik K, Cortese M, Healy BC, et al. Longitudinal analysis reveals high prevalence of Epstein-Barr virus associated with multiple sclerosis. Science. 2022;375(6578):296–301. 10.1126/science.abj8222

50. Lanz TV, Brewer RC, Ho PP, et al. Clonally expanded B cells in multiple sclerosis bind EBV EBNA1 and GlialCAM. Nature. 2022;603(7900):321–327. 10.1038/s41586-022-04432-7

## References

Tenca A, Jaakkola T, Farkkila M, et al. Environmental risk factors of pediatric-onset primary sclerosing cholangitis and autoimmune hepatitis. J Pediatr Gastroenterol Nutr. 2016;62(3):437–442.

Yasuda M, Yokode M, Tanaka A, et al. Anti-integrin alphavbeta6 autoantibodies in primary sclerosing cholangitis: a nationwide case-control study. J Gastroenterol. 2025.

Bloemen H, Mehandru S, et al. Anti-integrin alphavbeta6 antibodies in primary sclerosing cholangitis are associated with concomitant inflammatory bowel disease. Clin Gastroenterol Hepatol. 2025.

ElAbd H, et al. Convergent T-cell receptor repertoires and Epstein-Barr virus reactivity in primary sclerosing cholangitis. 2025.

Manfredo Vieira S, Hiltensperger M, Kumar V, et al. Translocation of a gut pathobiont drives autoimmunity in mice and humans. Science. 2018;359(6380):1156–1161.

Dorner H, et al. Outer membrane vesicles of PSC-associated pathobionts induce hepatic injury independently of bacterial translocation. 2024.

Burn L, Vyse A, Pilz A, et al. Incidence of Lyme borreliosis in Europe: a systematic review (2005-2020). Vector Borne Zoonotic Dis. 2023;23(4):172–194.

Xu Q, Kong Y, et al. Epidemiology of primary sclerosing cholangitis in China: a multicentre electronic medical record and literature-based study. J Autoimmun. 2024.

Lunder AK, Hov JR, Borthne A, et al. Prevalence of sclerosing cholangitis detected by magnetic resonance cholangiography in patients with long-term inflammatory bowel disease. Gastroenterology. 2016;151(4):660–669.

Bowlus CL, Arrive L, Bergquist A, et al. AASLD practice guidance on primary sclerosing cholangitis and cholangiocarcinoma. Hepatology. 2023;77(2):659–702.

Ancona S, Russell RK, et al. Oral vancomycin therapy in paediatric inflammatory bowel disease without primary sclerosing cholangitis. J Pediatr Gastroenterol Nutr. 2025.

Raisanen L, Lewindon PJ, et al. Oral vancomycin solution versus capsules in paediatric inflammatory bowel disease and sclerosing cholangitis. Br J Clin Pharmacol. 2025.

Wojciechowska-Koszko I, Kwiatkowski P, Sienkiewicz M, et al. Cross-reactive results in serological tests for borreliosis in patients with active viral infections. Pathogens. 2022;11(2):203.

## Supplementary References

51. Rich SN, Hinckley AF, Earley A, Petersen JM, Mead PS, Kugeler KJ. Tularemia, United States, 2011–2022. MMWR Morb Mortal Wkly Rep 2025;73(5152):1152–1156. 10.15585/mmwr.mm735152a1

52. Wildlife Informer. Deer population by state; accessed 20 April 2026, https://wildlifeinformer.com/deer-population-by-state/. See data file (Deer_density_provenance.docx), which will be released as described in the Data Availability Statement.

53. U.S. Department of Agriculture, Forest Service. Forest Inventory and Analysis (FIA) database: state forest cover, ca. 2016. St. Paul, MN: USDA Forest Service, Northern Research Station; accessed 2026. https://research.fs.usda.gov/programs/fia

54. Wieben CM. Estimated annual agricultural pesticide use by major crop or crop group for states of the conterminous United States, 1992–2017 (ver. 2.0). U.S. Geological Survey data release; 2019. 10.5066/P900FZ6Y

55. Dieter CA, Maupin MA, et al. Estimated use of water in the United States in 2015. U.S. Geological Survey Circular 1441; 2018. 10.3133/cir1441

56. U.S. Census Bureau. 2020 Census urban/rural classification; American Community Survey race and ethnicity estimates.

57. U.S. Environmental Protection Agency. Air Quality System (AQS) annual summary data, PM2.5 and ozone, 2018–2022. Research Triangle Park: EPA.

58. American Liver Foundation. Find a liver transplant center; accessed 2026. https://liverfoundation.org/

59. Centers for Disease Control and Prevention. Outpatient antibiotic prescriptions, United States, 2022 (IQVIA Xponent). Atlanta: CDC. https://arpsp.cdc.gov/resources/OAU-Data-Methods-2022.pdf

60. Centers for Disease Control and Prevention. Behavioral Risk Factor Surveillance System (BRFSS) age-adjusted prevalence, current smoker status, 2018–2022. Atlanta: CDC.

61. Grant CW, Juran BD, Ali AH, et al. Environmental chemicals and endogenous metabolites in bile of USA and Norway patients with primary sclerosing cholangitis. Exposome. 2023;3(1):osac011. 10.1093/exposome/osac011

62. Eisen L, Eisen RJ. Changes in the geographic distribution of the blacklegged tick, *Ixodes scapularis*, in the United States. Ticks Tick Borne Dis. 2023;14(6):102233. 10.1016/j.ttbdis.2023.102233

63. Bambha K, Kim WR, Talwalkar J, et al. Incidence, clinical spectrum, and outcomes of primary sclerosing cholangitis in a United States community. Gastroenterology. 2003;125(5):1364–1369. 10.1016/j.gastro.2003.07.011

64. Jaenson TGT, Jaenson DGE, Eisen L, et al. Changes in the geographical distribution and abundance of the tick *Ixodes ricinus* during the past 30 years in Sweden. Parasit Vectors. 2012;5:8. 10.1186/1756-3305-5-8 Lindkvist B, Benito de Valle M, Gullberg B, et al. Incidence and prevalence of primary sclerosing cholangitis in a defined adult population in Sweden. Hepatology. 2010;52(2):571–577. 10.1002/hep.23678

65. Schudel S, Gygax L, Kositz C, et al. Human granulocytotropic anaplasmosis – a systematic review and analysis of the literature. PLoS Negl Trop Dis. 2024;18(8):e0012313. 10.1371/journal.pntd.0012313

66. Dumler JS, et al. Human granulocytic anaplasmosis and *Anaplasma phagocytophilum*. Emerg Infect Dis. 2005;11(12):1828–1834. 10.3201/eid1112.050898

67. Karshima SN, Ahmed MI, Mohammed KM, Pam VA, Momoh-Abdullateef H, Gwimi BP. Worldwide meta-analysis on *Anaplasma phagocytophilum* infections in animal reservoirs: prevalence, distribution and reservoir diversity. Vet Parasitol Reg Stud Reports. 2023;38:100830. 10.1016/j.vprsr.2022.100830

68. Dahlgren FS, Heitman KN, Drexler NA, et al. Human granulocytic anaplasmosis in the United States from 2008 to 2012: a summary of national surveillance data. Am J Trop Med Hyg. 2015;93(1):66–72. 10.4269/ajtmh.15-0122

69. Kjær LJ, Klitgaard K, Soleng A, et al. Spatial patterns of pathogen prevalence in questing *Ixodes ricinus* nymphs in southern Scandinavia, 2016. Sci Rep. 2020;10(1):19376. 10.1038/s41598-020-76334-5

70. Quarsten H, Ryen CNB, Mørk LKT, et al. Tickborne *Neoehrlichia mikurensis* in the blood of blood donors, Norway, 2023. Emerg Infect Dis. 2025;31(11):2091–2097. 10.3201/eid3111.250125

71. Ponsioen CY, Defoer J, Ten Kate FJW, et al. A survey of infectious agents as risk factors for primary sclerosing cholangitis: are *Chlamydia* species involved? Eur J Gastroenterol Hepatol. 2002;14:641–648. 10.1097/00042737-200206000-00009

72. Hokynar K, Sormunen JJ, Vesterinen EJ, et al. *Chlamydia*-like organisms (CLOs) in Finnish *Ixodes ricinus* ticks and human skin. Microorganisms. 2016;4(3):28. 10.3390/microorganisms4030028

73. Pilloux L, Aeby S, Gaümann R, et al. The high prevalence and diversity of Chlamydiales DNA within *Ixodes ricinus* ticks suggest a role for ticks as reservoirs and vectors of *Chlamydia*-related bacteria. Appl Environ Microbiol. 2015;81(23):8177–8182. 10.1128/aem.02183-15

74. Kordick D, Brown T, Shin K, Breitschwerdt E. Clinical and pathologic evaluation of chronic *Bartonella henselae* or *Bartonella clarridgeiae* infection in cats. J Clin Microbiol. 1999;37:1536–1547. 10.1128/jcm.37.5.1536-1547.1999 Tenca A, Färkkilä M, Jalanko H, et al. Environmental risk factors of pediatric-onset primary sclerosing cholangitis and autoimmune hepatitis. J Pediatr Gastroenterol Nutr. 2016;62(3):437–442. 10.1097/mpg.0000000000000995

75. La Scola B, Holmberg M, Raoult D. Lack of *Bartonella* sp. in 167 *Ixodes ricinus* ticks collected in central Sweden. Scand J Infect Dis. 2004;36(4):305–306. 10.1080/00365540410020145

76. Bergh K, Bevanger L, Hanssen I, Løseth K. Low prevalence of *Bartonella henselae* infections in Norwegian domestic and feral cats. APMIS. 2002;110(4):309–314. 10.1034/j.1600-0463.2002.100405.x

77. Wass L, Grankvist A, Bell-Sakyi L, et al. Cultivation of the causative agent of human neoehrlichiosis from clinical isolates identifies vascular endothelium as a target of infection. Emerg Microbes Infect. 2019;8(1):413–425. 10.1080/22221751.2019.1584017

78. Höper L, Skoog E, Stenson M, et al. Vasculitis due to Candidatus *Neoehrlichia mikurensis*: a cohort study of 40 Swedish patients. Clin Infect Dis. 2021;73(7):e2372–e2378. 10.1093/cid/ciaa1217

79. Bloemen H, Livanos AE, Martins A, et al. Anti-integrin αvβ6 autoantibodies are increased in primary sclerosing cholangitis patients with concomitant inflammatory bowel disease and correlate with liver disease severity. Clin Gastroenterol Hepatol. 2025;23(9):1612–1622. 10.1016/j.cgh.2024.10.005

80. Yasuda M, Shiokawa M, Kuwada T, et al. Anti-integrin αvβ6 autoantibody in primary sclerosing cholangitis: a Japanese nationwide study. J Gastroenterol. 2025;60(1):118–126. 10.1007/s00535-024-02169-w

81. Roth D, Düll MM, Horst LJ, et al. Integrin αVβ6: autoantigen and driver of epithelial remodeling in colon and bile ducts in primary sclerosing cholangitis and inflammatory bowel disease. J Crohns Colitis. 2025;19(2):jjae131. 10.1093/ecco-jcc/jjae131

82. Yoshida H, Shiokawa M, Kuwada T, et al. Anti-integrin αvβ6 autoantibodies in patients with primary sclerosing cholangitis. J Gastroenterol. 2023;58(8):778–789. 10.1007/s00535-023-02006-6

83. Janse M, Lamberts LE, Franke L, et al. Three ulcerative colitis susceptibility loci are associated with primary sclerosing cholangitis and indicate a role for IL2, REL, and CARD9. Hepatology. 2011;53(6):1977–1985. 10.1002/hep.24307

84. Wakabayashi K, Lian ZX, Moritoki Y, et al. IL-2 receptor α−/− mice and the development of primary biliary cirrhosis. Hepatology. 2006;44(5):1240–1249. 10.1002/hep.21385

